# Flexibly encoded genome-wide association study identifies novel nonadditive genetic risk variants for cardiometabolic traits

**DOI:** 10.1101/2023.06.01.23290857

**Authors:** Jiayan Zhou, Andre Luis Garao Rico, Lindsay Guare, Million Veteran Program, Kyong-Mi Chang, Philip S. Tsao, Themistocles L. Assimes, Shefali Setia Verma, Molly Ann Hall

## Abstract

Most genome-wide association studies (GWAS) assume an additive inheritance model, which assigns heterozygous genotypes half the risk of homozygous-alternate genotypes. This has led to a focus on additive genetic effects in complex disease research. Growing evidence indicates that many single-nucleotide polymorphisms (SNPs) have nonadditive effects, including dominant and recessive effects, which are missed by the additive model alone. To address this issue, we developed Elastic Data-Driven Encoding (EDGE) to determine the inheritance model each SNP contributes to a given trait, allowing for unique and flexible SNP encoding in GWAS. Simulation results demonstrate that EDGE provides higher power than additive and other genetic encoding models across a wide range of simulated inheritance patterns while maintaining a conserved false positive rate. EDGE GWAS on data from the UK BioBank and the Million Veteran Program, comprising more than 500,000 individuals, identified nonadditive inheritance patterns for more than 52% of the genome-wide significant loci for coronary artery disease and body mass index. This research lays the groundwork for integrating nonadditive genetic effects into GWAS workflows to identify novel disease-risk SNPs, which may ultimately improve polygenic risk prediction in diverse populations and provide a springboard for future applications to thousands of disease phenotypes.

## Main

Although genome-wide association studies (GWAS) have identified hundreds of thousands of genotype–phenotype associations^1^, the majority of genetic variance for most complex diseases remains hidden. To analyze single-nucleotide polymorphisms (SNPs) in GWAS, the genotypes of each locus must be encoded with (i.e., turned into) meaningful numerical values. Most SNPs considered in GWAS have three possible genotypes: *AA* (homozygous for referent allele *A*), *Aa* (heterozygous), or *aa* (homozygous for alternate allele *a*). Traditional methods for encoding SNPs follow Mendelian inheritance models (dominant, recessive, or additive [incomplete dominant])^2,3^, which assume that *AA* genotypes have no risk (encoded score = 0) and *aa* genotypes have full risk (encoded score = 1). The assumed risk for *Aa* genotypes depends on the encoding method^4–6^. Dominant encoding assumes that heterozygous genotypes have full risk (encoded score = 1), whereas additive encoding assumes that heterozygous genotypes have half the risk of *aa* genotypes (encoded score = 0.5), and recessive encoding assumes that heterozygous genotypes have no risk (encoded score = 0). codominant encoding, which is a type of dummy encoding, allow for the possibility that *Aa* and/or *aa* genotypes have full risk. The number of encoding options reflects the wide range of possible SNP inheritance patterns, which vary within the genome among different loci and different phenotypes.

Since 2008, most GWAS have applied additive encoding^7–15^, limiting the discovery of SNPs with nonadditive inheritance patterns. Other encodings, including dominant and recessive, were used in the early days of GWAS along with additive encoding^16^, but additive encoding was soon adopted widely as a single approach to reduce the multiple hypothesis testing burden^4,6,17–19^. Not surprisingly, the greatest power to detect significant SNPs in association studies is achieved when the encoding matches the actual inheritance pattern of the SNPs^6^. Additive encoding alone is, therefore, underpowered to identify alleles with recessive effects^6^, even for common alleles^20^. Furthermore, the use of multiple traditional encoding approaches can limit the efficacy of GWAS, because an adjusted genome-wide significance threshold of 1.25 × 10^−8^ (genome-wide significance 5× 10^−8^/4) is needed to account for at least four tests for a single SNP, including all three Mendelian inheritance models and the codominant model. It is important, however, to identify the disease risks from recessive and dominant variants. For example, Manna *et al.*^21^ and Palmer *et al.*^22^ found many recessive and dominant effects in GWAS using UK BioBank data. Moreover, deleterious mutations tend to be recessive, as supported by both theory and empirical evidence. The recessive effects of some genetic variants might contribute significantly to phenotypic variation, alongside the many additive effects that have been identified through genome-wide association studies^21^.

Recently, alternative genetic encoding models have been established to incorporate additional genetic risks. For example, codominant encoding is a dummy encoding approach that allows heterozygous and homozygous-alternate genotypes to bear full risk in a single genetic model^23,24^, and dominance deviance (DOMDEV) is another common method in which the deviation of dominance from additivity is determined^22,25^. A drawback to the codominant encoding is that is does not estimate a single effect size per SNP for post-GWAS analyses, including meta-analysis and polygenic risk score calculation. Additionally, utilizing DOMDEV could increase the dimensionality of the genetic model, making it challenging to interpret the contributions of various genetic effects. This heightened complexity may also lead to potential issues with model overfitting. Growing evidence from studies that use flexible modeling suggests that the inheritance pattern of many SNPs is nonadditive, and these SNPs are often missed in GWAS when additive encoding is used alone^6,16,20,26^.

We previously developed Elastic Data-Driven Encoding (EDGE) as a flexible method for genetic encoding to help identify interactions among SNPs with nonadditive inheritance patterns^27^. EDGE allows unique, flexible, and data-informed encoding to be applied to each SNP. The large biobank datasets that are currently available offer the opportunity and power to identify SNPs with nonadditive inheritance patterns and large effect sizes. Here, we show that EDGE can accurately determine SNP inheritance models and outperform additive and other encoding methods when applied in large-scale simulated GWAS. Furthermore, in GWAS with more than a half million participants from the UK BioBank and the Million Veteran Program (MVP), EDGE identified both additive and novel nonadditive SNPs associated with coronary artery disease (CAD) and body mass index (BMI). Our results demonstrate that flexible encoding based on the unique inheritance patterns of SNPs enables the detection of SNP–trait associations beyond the additive inheritance model in GWAS.

## Methods

### Elastic Data-Driven Encoding

As previously described^27^, EDGE, like other encodings, encodes the *AA* and *aa* genotypes with values of 0 and 1, respectively. In contrast to other encodings, EDGE assigns the heterozygous genotype (*Aa*) for each SNP a flexible, heterozygous–to–homozygous-alternate risk ratio, designated as α, based on the inheritance pattern of the SNP. For example, if the underlying genetic inheritance pattern of SNP *A* is additive, then genotype *Aa* has 50% the risk of genotype *aa*, and EDGE assigns the heterozygous genotype an α value of 0.5. The value of α is calculated by the following equations:

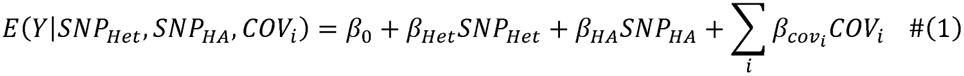

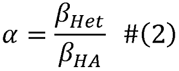

where *E(Y)* is the expected value of the given phenotype (*Y*), *SNP_HET_* is the heterozygous encoding, *SNP_HA_* is the homozygous alternate encoding, *β_Het_* and *β_HA_* are the regression coefficients for the heterozygous and homozygous alternate encodings, respectively, *COV_i_* is the *i*th covariate, and *β_COVi_* is the corresponding regression coefficient.

### Simulated datasets

To ensure that EDGE assigns the expected α values across SNPs with different underlying inheritance patterns, we simulated main effect SNPs with the following genetic inheritance patterns using the Biallelic Model Simulator (BAMS)^27^ within pandas-genomics Version 0.11.0: null or random effect, recessive, sub-additive, additive, super-additive, and dominant. Simulations were performed with 1,000 simulated datasets for each inheritance pattern across varying minor allele frequencies (MAFs; **Fig. 1**). In each simulation, one SNP demonstrated a main effect. To avoid overfitting, each simulated dataset consisted of two separate subsets: one to calculate α values (EDGE α calculation dataset) and one to apply the α values to obtain p-values and effect estimates (GWAS dataset). Other encodings, including additive, recessive, dominant, codominant, and DOMDEV, were also applied to the second subsets for p-value and power comparisons. Key parameters were considered in order to exemplify possible data structures in natural biomedical datasets, including MAF = 0.05, 0.1, 0.2, 0.3, and 0.4; sample size = 2,000, 5,000, 10,000, 50,000, and 100,000; case-control ratio = 1:1 and 1:3; and penetrance difference (the difference between the minimum and maximum probabilities in the penetrance table) = 0.05, 0.1, 0.175, 0.25, 0.33, and 0.4, as shown in **Fig. 1**.

**Fig. 1.**
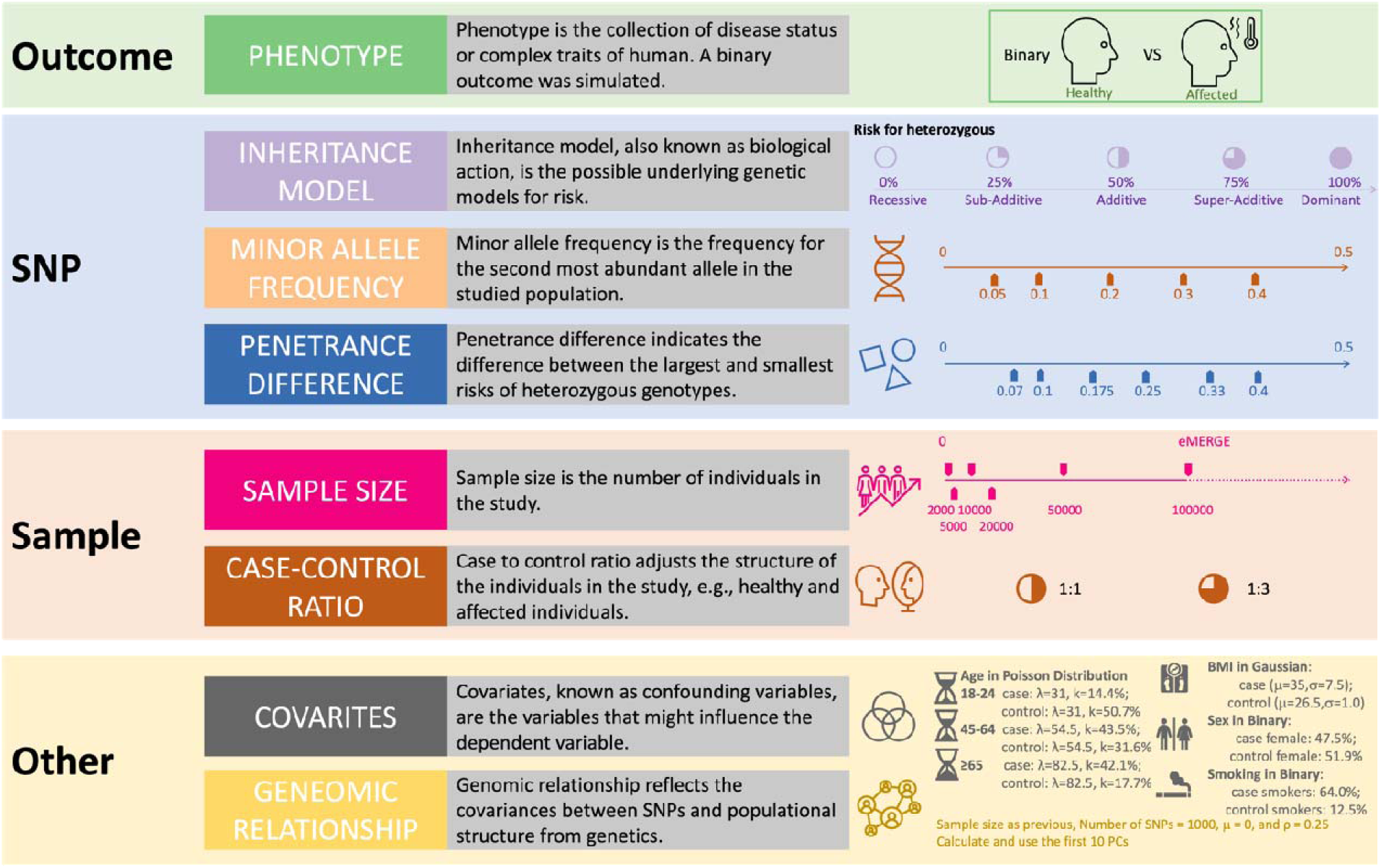
Workflow of the simulation study to compare the performance of EDGE to traditional encoding methods.

In addition, we simulated two continuous confounder variables and two binary variables for cases and controls separately to assess their potential impacts. The simulated covariates and distributions were referenced from the National Diabetes Statistics Report, 2020^28^. Age (years) was simulated using the Poisson distribution for three different groups, including 18 to 44 (case: λ = 31, k = 14.4%; control: λ = 31, k = 50.7%), 45 to 64 (case: λ = 54.5, k = 43.5%; control: λ = 54.5, k = 31.6%), and ≥ 65 (case: λ = 82.5, k = 42.1%; control: λ = 82.5, k = 17.7%). BMI was simulated by considering the Gaussian distribution for cases (μ = 35, σ = 7.5) and controls (μ = 26.5, σ = 1.0). Two binary covariates were generated by referencing the binomial distributions of sex (case female = 47.5%; control female = 51.9%) and smoking status (case smokers = 64.0%; control smokers = 12.5%).

To presume the population structure derived from the genotyping data for each simulation, we additionally simulated a genomic relationship matrix (G-matrix) using a variance-covariance matrix with all covariance at 0.25 for 1,000 SNPs and the corresponding sample size. We then performed a principal component analysis (PCA) and used the first 10 principal components (PCs) as covariates to represent the relationships between individuals.

To further evaluate the performance of EDGE across a spectrum of variant frequencies, we performed simulations with 1,000 replicates using lower MAFs (0.025, 0.01, 0.005, and 0.001) and sample sizes (from 2,000 to 100,000) with a case:control ratio of 1:3, four preset covariates (age, sex, BMI, and smoking status), and six inheritance patterns (recessive, sub-additive, additive, super-additive, dominant, and heterozygous). The non-convergence rate was calculated as the ratio between the number of models that failed to converge and the total number of replicates that could be finished.

### Single-variant association tests

We conducted single-variant association tests with a simulated binary outcome using logistic regression for simulated datasets encoded using EDGE and the other traditional encoding schemes separately. The EDGE α calculation datasets used to calculate α for EDGE were simulated using the same parameters as the datasets used to obtain the corresponding p-value and effect estimates, but with different sets of seeds to control the randomness. Another same set of simulated GWAS datasets, different from the EDGE α calculation datasets, was used to assess p-values and effect estimates for EDGE and the other encodings, allowing for a direct comparison of their performance. Three different schemes were applied to the single-variant association tests, including models that included SNP only, SNP with the four covariates, and SNP with the four covariates and the first 10 presumable PCs. The power was obtained for each combination of parameters. We used a genome-wide significance threshold (5 × 10^−8^) to assess the encoding methods for their power at the GWAS level. The false positive rate was calculated from the simulation with null effect SNPs for every combination of encodings and parameters.

### Comparing power and α calculation across parameters

Pairwise comparisons were performed to compare the power of each encoding method. We considered the simulated data with a penetrance difference of 0.1 and sample size from 5,000 to 50,000 with all possible MAFs, case-control ratios, and inheritance models as a representation of the majority of GWAS that have been performed. Random forest analyses were conducted to rank the importance of parameters to the power and α calculation using the *RandomForest* package in R v4.2.0. Parameter importance was computed using the percentage increase in mean squared error and was expressed relative to the maximum.

### Alpha calculation and EDGE GWAS in UK BioBank

UK BioBank is a large-scale prospective cohort study that focuses on health-related outcomes for over 500,000 recruited participants through surveys, direct physical measurements, and analyses of biological specimens^29^. We applied the EDGE algorithm to the UK BioBank data (project #52374) for CAD as a binary outcome and BMI as a continuous outcome. Participants with CAD were identified using standardized clinical classifications, including 9^th^ revision of the International Classification of Diseases (ICD-9) codes (410–412), ICD-10 codes (I210–I214, I219–I221, I228, I229, I231–I233, I236, I214, I240, I241, I248, I249, and I252), and OPCS-4 codes (K40–K46, K49, K501, and K75), and additional self-reported or doctor-reported status, including doctor-diagnosed vascular heart problems (1: Heart attack; 2: Angina), non-cancer illness codes (1074: Angina; 1075: heart attack/myocardial infarction), and operation codes (1095: coronary artery bypass graft; 1523: triple heart bypass; 1070: coronary angioplasty +/- stent)^30,31^. Individuals who did not meet any of the above classifications were considered controls. For analyses with BMI as the outcome, we used the same population used for CAD but only included participants with a documented BMI at enrollment. BMI was log10-transformed to adjust for the high skewness.

Individuals of European (EUR; BMI: 313,322; CAD: 314,998 with 27,310 cases and 287,688 controls) or African (AFR; BMI: 9,504; CAD: 9,636 with 560 cases and 9,076 controls) descent were used for ancestry-specific EDGE GWAS. The PCs for the EUR and AFR parameters were re-calculated independently using PC-AiR^32^ after removing the 3^rd^-degree relatives based on KING^33^. The TOPMed imputed hard call genotypes were obtained for each ancestry group by chromosome separately. Individuals were excluded if they had more than 1% missing genotypes. SNPs were excluded if they met any of the following criteria: more than 1% missing genotypes, MAF less than 1%, Hardy-Weinberg equilibrium test p-value below 1 × 10^−6^ with mid-p adjustment, or imputation R^2^ < 0.3. For each ancestry group, the data were split randomly into two sets of equal size, one to calculate α using EDGE and the other to apply the α in an EDGE GWAS, while maintaining the same distribution (case-control ratio for CAD and normal distribution for log10(BMI)). The dataset used for EDGE α calculation was used to generate the α value for each SNP (using Equation 1 and Equation 2). Specifically, heterozygous genotypes were encoded using the calculated α values, and further GWAS analyses were conducted based on the EDGE encoding (homozygous referent = 0, heterozygous = α, homozygous alternate = 1) using the EDGE GWAS dataset. Sex, age at enrollment, genotyping batch number, and the first 10 PCs were adjusted as covariates.

### EDGE GWAS in Million Veteran Program

The Veterans Health Administration (VA) has recruited over 1,000,000 active participants since 2011 from more than 75 VA Medical Centers nationwide, and the study design of the MVP has been previously described^35^. Every participant has provided informed consent along with biological samples and access to their full electronic health record within the VA system before and after enrollment. This study (MVP003/028) was approved by the VA Central Institutional Review Board for ethical and protocol compliance.

Participants with CAD were identified based on ICD9 codes (410–414) with a requirement of at least two occurrences in separate encounters. Additionally, participants who had undergone at least one coronary angiogram were included. The detailed classification of CAD cases within MVP was described previously^8^. BMI at enrollment was extracted for EDGE GWAS analysis. Both CAD and BMI were studied with sex and the first 10 PCs as covariates. The ancestry of each participant was determined using Harmonizing Genetic Ancestry and Self-Identified Race/Ethnicity (HARE)^36^, a procedure that combines self-identified ethnicity and genetically inferred ancestry. EDGE GWAS were conducted exclusively with individuals identified as having European ethnicity by HARE. Other ethnic groups were not included because of concerns about sample size and statistical power. In these EDGE GWAS, SNPs were encoding using α values calculated from the UK BioBank for EUR CAD (453,120 with 142,596 cases and 310,524 controls) and EUR BMI (391,659), respectively.

### Meta-analysis of the EDGE GWAS outputs

We conducted meta-analyses for each of the encodings with the summary statistics from the EDGE GWAS, incorporating data from both the UK BioBank and the MVP for both phenotypes. The meta-analysis was performed using PLINK 1.9^34^ for both fixed and random effects. We excluded SNPs with stringent linkage disequilibrium (LD) criteria of r^2^ = 0.8 within 500 kb and I^2^ ≤ 40 during the post-GWAS quality control. To recover potentially relevant associations that may have been filtered out due to LD, we subsequently used the LDtrait tool (NIH LDlinkR API using R)^37^ using a more relaxed linkage disequilibrium threshold of r^2^ = 0.1 within 500 kb. This allowed us to correlate our significant variants with traits and diseases previously reported in the GWAS Catalog. Novel GWAS signals were obtained by excluding known SNPs associated with the two outcomes and their related phenotypes, such as height and weight for BMI and a list of heart diseases for CAD, including cardiovascular disease, coronary heart disease, myocardial infarction, atrial fibrillation, hypertrophic cardiomyopathy, and ischemic stroke. We plotted the distribution of α values against the MAF for significant SNPs and generated Manhattan plots and Q-Q plots using R version 4.3.2.

## Results

### EDGE detected common and rare variants for simulated SNPs across a range of inheritance models

A total of 2.1 million SNPs were simulated in the study to assess the performance of EDGE compared to traditional encodings. The density distributions of α peaks corresponded to the respective simulated inheritance models (recessive ≅ 0, sub-additive ≅ 0.25, additive ≅ 0.5, super-additive ≅ 0.75, and dominant ≅ 1; **Fig. 2a**), demonstrating the efficacy of α to infer inheritance models for additive and nonadditive SNPs. Next, we contrasted the average power and significance of each encoding method for SNPs with distinct inheritance patterns, using 50,000 samples with MAF = 30% and a 1:3 case:control ratio (1,000 datasets for each inheritance model) as an example (**Fig. 2b**). The MAF of 30% was selected by referencing the first reported associations for type 2 diabetes with the E23K variant in *KCNJ11* (MAF ∼ 30% for Europeans)^38^. EDGE detected signals with genome-wide significance (5 × 10^−8^) for more inheritance patterns than any of the other encoding methods, and it was the top-performing method for every inheritance pattern. No method identified SNPs with recessive inheritance; however, EDGE identified SNPs with all other inheritance patterns, unlike any other encoding method, including additive encoding. Codominant encoding performed nearly as well as EDGE; however, our previous study showed that codominant encoding suffered from model non-convergence for lower MAFs^27^. Regarding inflation, EDGE demonstrated a conserved false positive rate of 5.4%, which falls within Bradley’s liberal criteria of 2.5–7.5%^39^ (**Fig. S1**).

**Fig. 2.**
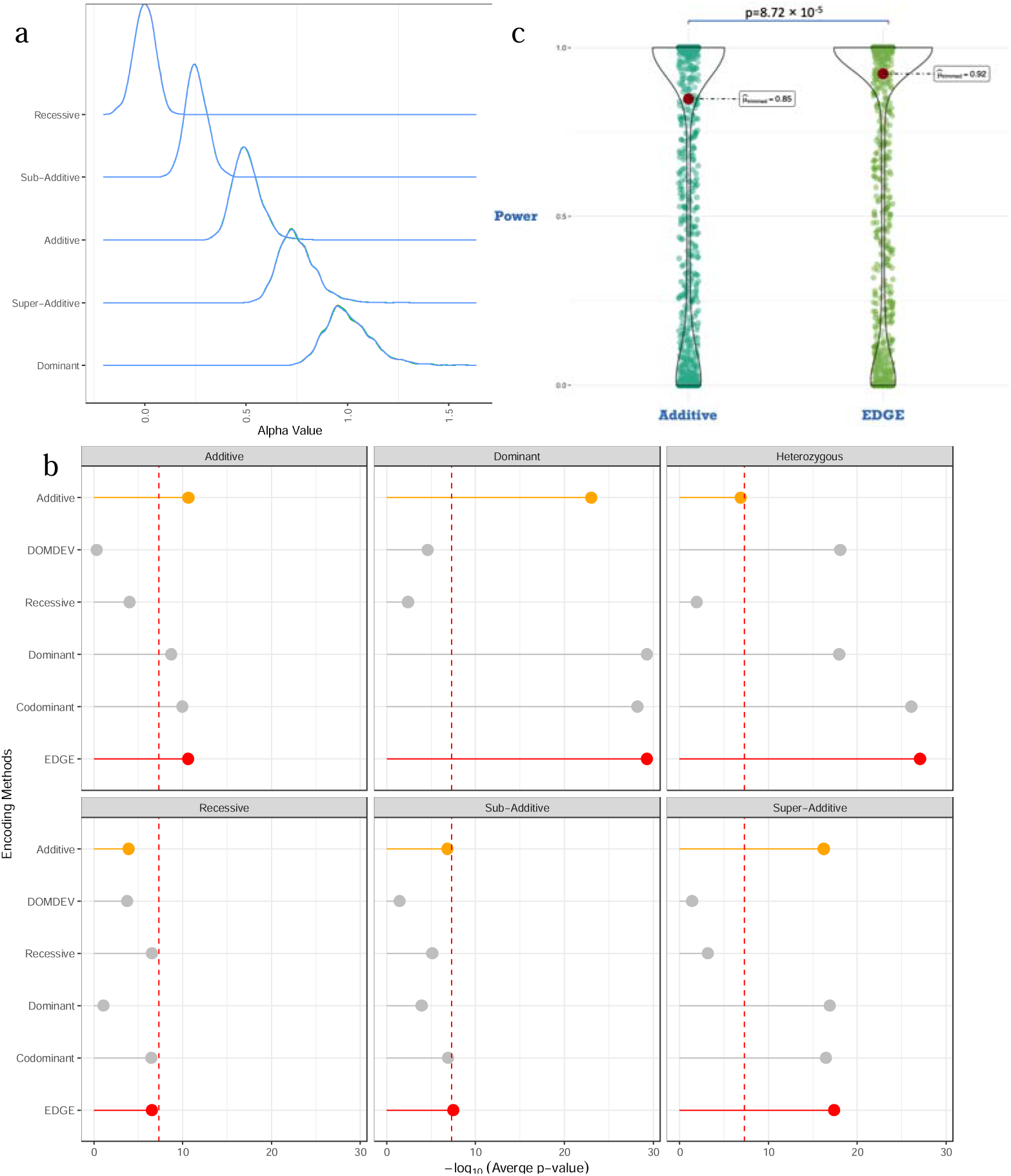
EDGE performance in the simulations. a) Distribution of α values for SNPs with different inheritance models. b) SNPs with MAF = 30% were simulated for 12,500 cases and 37,500 controls. The red dashed line represents the significance threshold at the genome-wide scale (5 × 10^−8^). c) Power comparison between additive encoding and EDGE considering all possible inheritance models.

To ensure the robustness of EDGE across the parameters of interest, we extended our simulation to consider all the possible parameters indicated in **Fig. 1**. We observed that the power increased with sample size and MAF. The EDGE and codominant encoding could achieve the desired power of 80% more steadily with increasing sample size than the other traditional encodings for SNPs with the lowest MAF of 0.05 (**Fig. 3a**). We compared the difference in median power across all combinations of parameters between EDGE and additive encoding using Kruskal– Wallis χ^2^ tests. EDGE had significantly more power than additive encoding (p = 8.7 × 10^−5^; **Fig. 2c**). Of note, EDGE identified SNPs with recessive inheritance and heterozygous inheritance that additive encoding failed to detect (**Fig. 3b**). Each of the other traditional encodings lacked sufficient power to detect SNPs with one or more of the inheritance patterns. For example, the dominant and DOMDEV encodings failed to detect SNPs with recessive or sub-additive inheritance with limited samples. EDGE had robust power to identify SNPs under any inheritance pattern, even the sub-additive and recessive patterns that other encoding failed to detect with sufficient power. With low MAF (0.05) and small sample size, EDGE demonstrated greater power than the other encodings to capture associations between SNPs and the outcome. For example, additive encoding required more than 10,000 samples to attain the desired power (80%) for SNPs with low MAF.

**Fig. 3.**
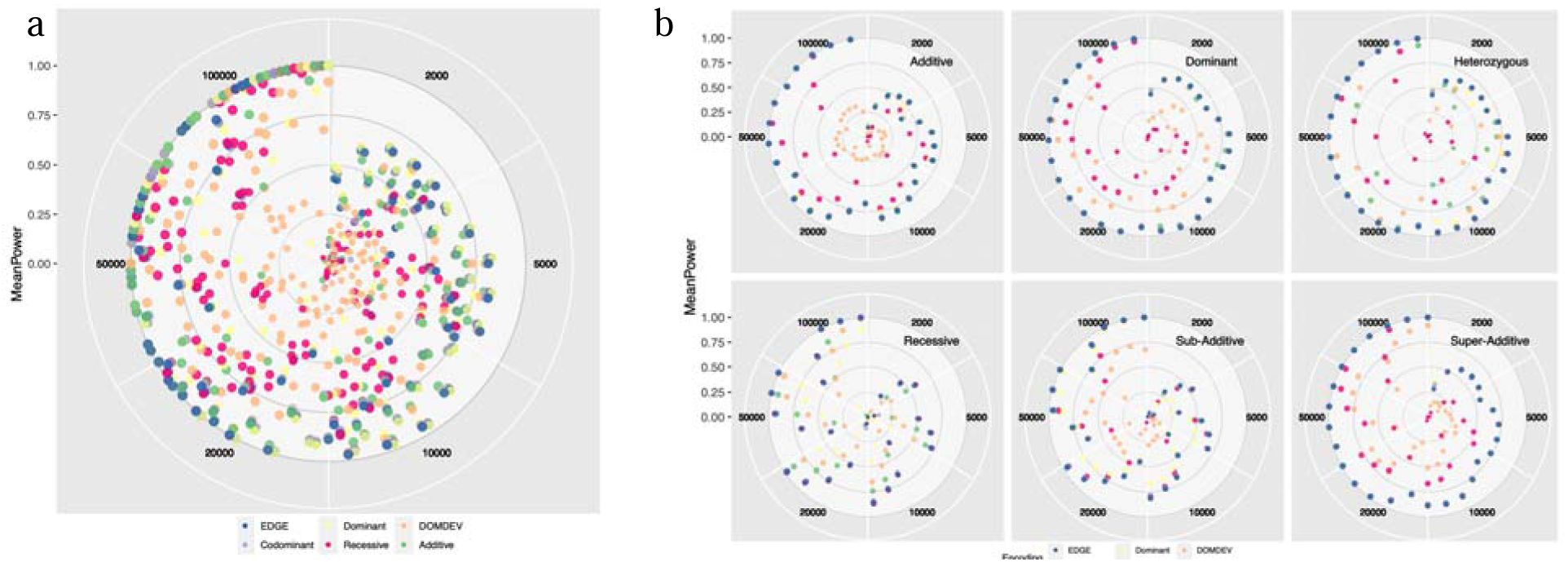
The power of EDGE and the other encodings to detect SNP associations a) across all simulated models and parameters and b) by inheritance patterns. The sample size increases from 2,000 to 10,0000 clockwise. In each sample size segment, the MAF increases left to right from 0.05 to 0.4. The power increases from the center of the plot outward.

Because all the encodings were challenged by simulated scenarios where there were few or no individuals with *aa* genotypes, which likely led to reduced minor allele counts (MAC), we estimated the convergence percentage for using EDGE in regressions for SNPs as common and rare variants, varying the MAF and sample size. The convergence rate for EDGE was 99.35% when testing common SNPs with a low MAF (0.05) and a sample size of 2,000 (**Fig. S2**). We then extended our simulations to include SNPs with progressively lower MAFs of 0.025, 0.01, 0.005, and 0.001 across various sample sizes while incorporating covariates. The convergence rate for EDGE dropped below 50% in simulations with 2,000 samples when the MAF fell below 0.01; however, we observed that increasing the sample size could potentially mitigate the convergence issues when applying EDGE to less frequent genetic variants.

### EDGE ensured stable α calculations and power across models, highlighting key factors in single variant association analysis

We assessed the ability of EDGE to accurately detect significant variants with a small sample size and low MAF, which were two factors that significantly influenced the power and α calculations in the parameter importance tests (**Fig. S3**). We constructed three models to test the stability of the α calculation and power for EDGE, including SNP, SNP with covariates, and SNP with covariates and PCs as the population structure. We then executed two overfitted random forest paradigms to uncover the importance of variables to the power and α calculation. The results showed that sample size and penetrance had the greatest importance for the power, followed by the encoding method, the SNP inheritance pattern, and the MAF (**Fig. S3a**). The α calculation was affected by the MAF but not by the inheritance pattern (**Fig. S3b**). Neither calculation was altered by the case:control ratio or the covariates and PCs.

EDGE was able to identify more significant results than the other encodings at the same combination of MAF and sample size, particularly for SNPs with heterozygous, dominant, or super-additive inheritance patterns (**Fig. S4**). Codominant encoding functioned similarly to EDGE, as both demonstrated relatively high statistical power and yielded significant results reflected in small p-values. Therefore, we restricted the analysis to codominant encoding and EDGE to compare the significance of the results. EDGE was more likely than codominant encoding to reveal significant results with the smallest MAF (0.05, Kruskal–Wallis χ^2^ = 34.24, p = 4.87 × 10^−9^) and smallest sample size (2000, Kruskal–Wallis χ^2^ = 72.27, p = 1.88 × 10^−17^; **Table S1**). We also compared the performance of EDGE and codominant encoding using simulations incorporating covariates and PCs. We found that EDGE consistently assigned more significance in simulations with smaller MAFs and sample sizes.

We next evaluated the distribution of α values under different combinations of MAF, penetrance difference, sample size, and case:control ratio. The estimation of α values for single SNPs was steady regardless of the number and types of covariates and population structures. We observed distinct density peaks that aligned with the simulated inheritance patterns, signifying that the α values from EDGE reflect the SNP inheritance patterns (**Fig. S5**). As the MAF, sample size, and baseline risk increased, the density peaks became sharper, converging on the values corresponding to the SNP inheritance patterns and showing greater separation among different inheritance patterns. Conversely, decreases in sample size, MAF, and baseline risk caused the distribution of α values to broaden.

### EDGE GWAS identified novel additive and nonadditive significant findings for CAD and BMI using UK BioBank and MVP

The EDGE algorithm comprises two primary steps: 1) calculate α values by considering the effect of heterozygous genotypes relative to homozygous alternate genotypes and 2) apply these SNP-specific α values in EDGE GWAS using regression, where they are used to encode heterozygous genotypes. We computed α values for each SNP using half the UK BioBank participants for binary (CAD) and continuous (BMI) outcomes within the European and African ancestry cohorts (**Table S2**). Subsequently, we applied the α values to encode heterozygous effects in EDGE GWAS using the remaining UK BioBank participants as a single-variant test for each phenotype. To further assess the resulting alpha values in the Mendelian inheritance pattern categories, we manually assigned α value ranges to each of the inheritance models as follows: recessive (α = 0 to 0.125), sub-additive (α = 0.125 to 0.375), additive (α = 0.375 to 0.625), super-additive (α = 0.625 to 0.875), dominant (α = 0.875 to 1), and two additional categories for under-recessive (α < 0) and over-dominant (α > 1).

The individual GWAS results are available in the Supplementary Information (UK BioBank: **Figs. S6–S9 and Tables S3–S6**; MVP: **Figs. S10–S11**). We did not observe major genetic inflation within GWAS analyses, as depicted in **Figs. S12–S13**. We performed meta-analyses for BMI (**Fig. 4a and Table S7**) and CAD (**Fig. 4b and Table S8**) by integrating the EDGE GWAS results from the UK BioBank and MVP cohorts. The top results for CAD included SNPs near or within *CDKN2B-AS1*/*CDKN2A* (9p21 region, α = 0.23 to 0.69; p = 5.20×10^-159^ to 4.38×10^-10^), *LPA* (α = 0.65; p = 1.57×10^-150^), *LDLR* (α = 0.47; p = 1.03×10^-32^) and *APOE* (α = 0.69; p = 3.17×10^-22^). More than 50% (57.65%) of the significant findings exhibited nonadditive inheritance patterns, such as SNPs within *ABCG8* (α = 0.09; p = 4.21×10^-8^) and *AP002989.1* (α = 0.7; p = 4.27×10^-19^). The top results for BMI included SNPs near or within *FTO* (α = 0.36 to 0.62; p = 0 to 2.60×10^-8^), *TMRM18* (α = 0.59; p = 5.68×10^-91^), *SEC16B*/*CRYZL2P-SEC16B* (α = 0.22 to 0.64; p = 3.76×10^-19^ to 3.85×10^-8^), and *TCF7L2* (α = 0.25 to 1.28; p = 3.96×10^-44^ to 7.89×10^-9^). Multiple significant SNPs (52.4%) exhibited nonadditive inheritance patterns, such as SNPs within *LINC01874* (α = 0.92; p = 7.02×10^-57^) and *RPS17P5* (α = 1.06; p = 2.80×10^-33^). We also observed that EDGE could identify significant SNPs with smaller effects (**Fig. S14 and Fig. S15**) on both types of phenotypes (binary and quantitative). SNPs with very small allele frequencies had large effects, with a negative Pearson’s product-moment correlation (–0.1146; p = 0.0003).

**Fig. 4.**
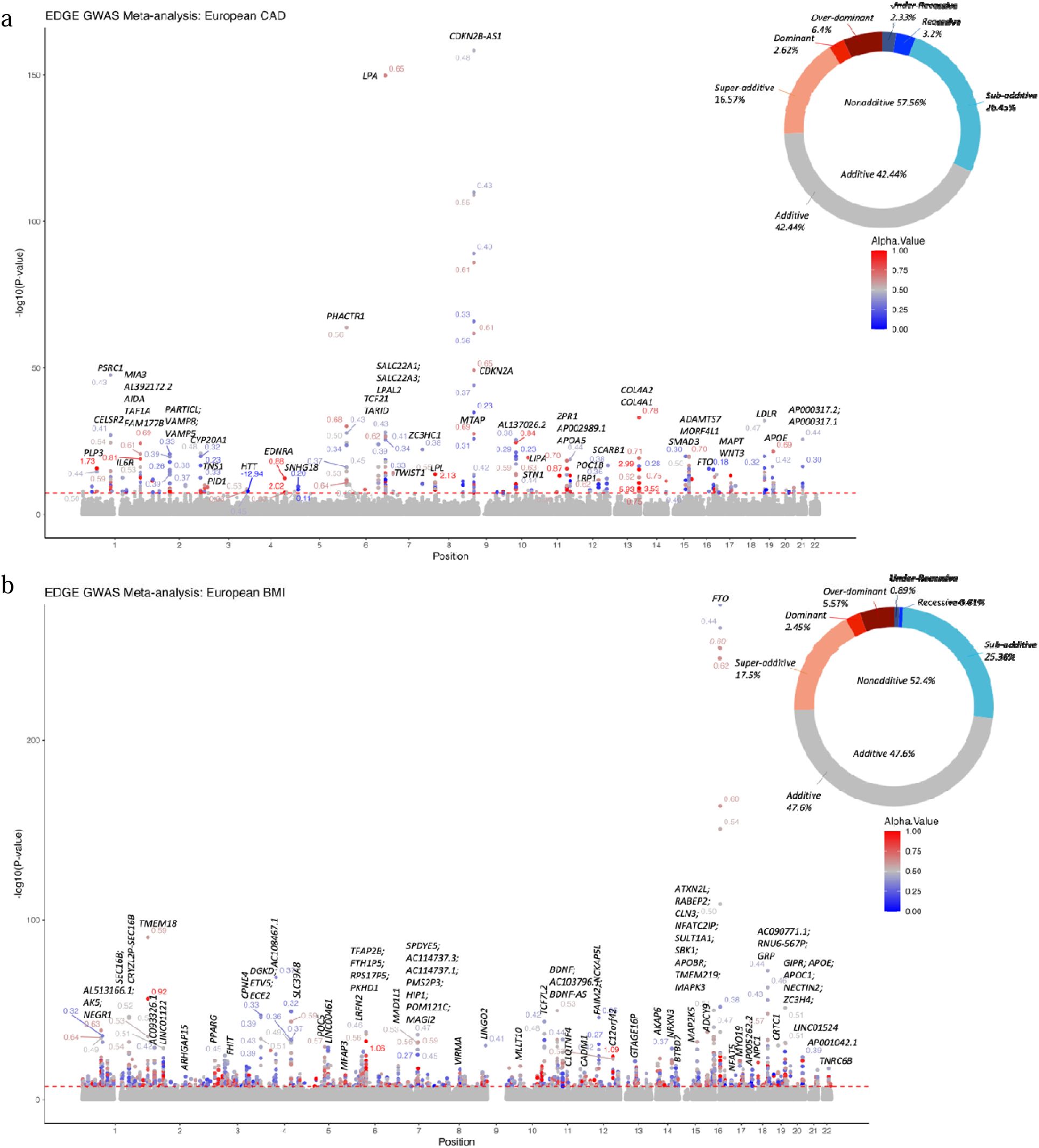
Meta-analysis for SNPs contributing to a) CAD and b) BMI in the European ancestry group. The red dashed line represents the genome-wide significance threshold at 5 × 10^−8^. The percentages on the circles represent the proportion of each inheritance pattern for each phenotype: recessive (α < 0.125), sub-additive (α = 0.125 to 0.3755), additive (α = 0.375 to 0.625), super-additive (α = 0.625 to 0.875), an dominant (α > 0.875).

To assess EDGE’s ability to identify previously reported loci, we used the LDtrait tool to perform lookups in the GWAS Catalog for the significant hits in our meta-analyses, applying a linkage disequilibrium threshold of r^2^ = 0.1 in a 500 kb window. We then removed all SNPs with known phenotype associations to identify novel associations between SNPs and the phenotypes, such as SNPs in 9p21 region, *LPA*, *LDLR*, *APOB* and *APOE* for BMI, and SNPs in *FTO* for BMI.

We identified four previously unreported loci associated with BMI (**Table S9**), including rs12881530 (intergenic, α = 0.59, beta = –0.0005, MAF = 0.406) previously known to be associated with smoking status, red cell width, insomnia, and response to paliperidone in schizophrenia; rs35420030 (*FTO*, α = 0.43, beta = –0.0002, MAF = 0.491) associated with lung function; rs7594041 (intergenic, α = 0.46, beta = 0.0007, MAF = 0.406) associated with smoking status, total and LDL cholesterol levels, and brain function; and rs116717000 (intergenic, α = 2.57, beta = –0.006, MAF = 0.050) associated with effects of food intake, especially protein, fat, and vegetables, on BMI.

EDGE discovered 15 novel loci associated with CAD (**Table S10**), including rs28557075 (α = 0.32, OR = 0.9693, MAF = 0.078) and rs62560775 (α = 0.34, OR = 0.9748, MAF = 0.091), both within the 9p21 region (*CDKN2B-AS1*) and associated with multiple cancers; rs783147 (*PLG*, α = 0.43, OR = 0.9852, MAF = 0.444) and rs72925025 (BMPR2, α = 0.33, OR = 1.0251, MAF = 0.156) associated with levels of total, LDL, and HDL cholesterol (a) and other lipids; rs6010690 (*TPD52L2*, α = 0.50, OR = 1.0122, MAF = 0.405) and rs79965102 (*TRPC4AP*, α = 0.47, OR = 1.028, MAF = 0.050) associated with BMI and blood pressure; rs17881561 (*FCSK*, α = 0.14, OR = 0.944, MAF = 0.059) associated with red blood cell features, blood pressure, and cholesterol levels; and multiple intergenic SNPs, including rs74608880 (α = 0.25, OR = 1.0605, MAF = 0.049), rs386453 (α = 1.63, OR = 1.0073, MAF = 0.107), rs1611873 (α = 0.744, OR = 0.9889, MAF = 0.447), rs540713, rs1247562 (α = 0.877, OR = 1.0106, MAF = 0.152), rs115682464 (α = –12.94, OR = 1.001, MAF = 0.057), rs62163129 (α = 1.308, OR = 1.0083, MAF = 0.080), and rs73023864 (α = 44, OR = 1.0154, MAF = 0.189) associated with cancers, type 2 diabetes, BMI, blood pressure, red blood cell measurements, and lipid levels.

According to the EDGE α values, less than half the significant results in the meta-analyses had additive inheritance (47.60% [854/1794] for BMI; 42.44% [146/344] for CAD; **Fig. 4**). Among novel significant SNPs, deviations from additive inheritance were common, accounting for 67% (10/15) of CAD SNPs and 25% (1/4) of BMI SNPs. Importantly, our analysis revealed a notable relationship among the inheritance model, minor allele frequency (MAF), and the effect size (log(OR)).As shown in **Figs. S14 and S15**, SNPs with low MAF tend to exhibit recessive effects, corresponding to high log(OR) values. This pattern was consistent across both CAD and BMI analyses.

## Discussion

In this study, we introduced EDGE, a SNP encoding method for GWAS that flexibly represents each SNP based on its inheritance for a given phenotype. This approach enhances the identification of novel, nonadditive SNPs. EDGE is particularly effective in uncovering the inheritance patterns underlying SNP–disease associations, even when the inheritance models are not initially known. SNPs with nonadditive inheritance can be overlooked by traditional additive encoding because of their complex genetic architecture. By employing a multi-encoding EDGE GWAS approach, we were able to maintain conservative analytical power without the risk of multiple-test corrections rendering the results insignificant.

Extensive simulations showed that EDGE is capable of 1) assigning α values in line with the simulated SNPs’ inheritance model, and 2) identifying both additive and nonadditive variants contributing to binary and continuous outcomes. EDGE retained higher analysis power and the ability to detect significant signals, even with an extremely small sample size (2,000) or a MAF (0.05). EDGE also exhibited a low false positive rate, showcasing its reliability and accuracy in genetic association studies, similar to those of the additive encoding commonly used in the field.

We applied the EDGE algorithm to calculate α values using half the available UK BioBank data. We then conducted EDGE GWAS using the calculated α values on the remaining UK BioBank data and the MVP data. Finally, we conducted a meta-analysis combining the data from both sources. The most common genetic risk factors with additive inheritance were successfully replicated by EDGE GWAS, including variants in *FTO* (α = 0.36 to 0.62) for BMI and variants in the 9p21 region (*CDKN2B-AS1*/*CDK2NA*; α = 0.33 to 0.65) for CAD. Novel variants were also identified for both BMI and CAD. Some new variants within genes with previously reported phenotypic associations were newly associated with the same phenotypes, including rs35420030 near the *FTO* gene associated with BMI, and rs28557075 and rs62560775 near the *CDKN2B- AS1* gene in the 9p21 region associated with CAD. EDGE GWAS also identified additional variants near other genes or within intergenic regions. These newly discovered variants have been previously associated with traits that might have direct or indirect effects (i.e., as a mediator) on BMI or CAD, suggesting potential biological hypotheses for further analyses.

EDGE GWAS offers a powerful approach to assign α values (as indicators for inheritance patterns) based on available data, allowing associations between risk variants and traits to be studied under both additive and nonadditive inheritance models simultaneously. Unlike traditional GWAS, which primarily focus on additive effects, EDGE GWAS can identify significant risk variants while also uncovering opportunities for much larger effect sizes with smaller p-values than are typically found for alleles with additive inheritance. Most GWAS tend to focus on additive effects, often missing SNPs with recessive effects, particularly those with low MAF. By capturing these recessive effects, our method has increased power to identify significant SNP–disease associations. This is crucial, as it allows us to uncover genetic variants with substantial impacts that have been missed by traditional approaches. EDGE GWAS thus offers an alternative approach to conducting GWAS with conservative and comparable power, maintaining a low false positive rate by employing a data-driven, flexible encoding strategy instead of uniformly applying additive or dominant encoding for every SNP.

Our research has several limitations. The EDGE GWAS algorithm relies on α values to recode heterozygous genotypes, which must be generated using either a portion of the dataset or the entire dataset. The use of the same data to both calculate and apply α values will lead to significant inflation of the false positive rate. Conversely, if the dataset is split to maintain a low false positive rate, analytic power may be lost. A potential solution might involve leveraging a large cohort, such as UK BioBank or MVP, to generate α values for SNPs and then distributing the calculated α values as a universal α directory along with summary statistics. Other study groups would then be able to apply the α values to non-overlapping participants in different cohorts. In addition, the regression model suffered from poor fitting due to a lack of convergence for low-frequency variants (MAF < 0.001) with few participants. However, EDGE provides a robust capability to identify common variants in the GWAS setting with an extremely small sample size. Second, we found that the sample size was suboptimal for smaller populations in the studies (East Asian, South Asian, and Admixed American). Consolidation of other available datasets with diverse ancestry is needed to enable better genetic analyses of diseases within different ancestry subgroups. Our current strategy for ancestry-based stratification combines individuals with Admixed American ancestry into one dataset and uses PCs as covariates. This might not reflect the diversity and richness of genetic risk profiles in this population. Therefore, we need additional approaches to overcome this limitation and include the full spectrum of diversity in our research.

In conclusion, EDGE effectively identifies both additive and nonadditive SNPs as implicated by simulation studies and association studies for CAD and BMI. Through its novel computational framework and inheritance pattern estimation capabilities, EDGE overcomes the limitations of traditional additive-only models in GWAS. This flexibility allows researchers to detect SNPs with dominant, recessive, and intermediate inheritance patterns that would otherwise be missed. EDGE’s approach directly improves the accuracy of gene-environment interaction studies and polygenic risk scores by incorporating the true genetic architecture rather than assuming additivity for all variants. This leads to more precise complex trait and disease prediction models that better reflect biological reality. EDGE provides a more comprehensive understanding of the genetic architecture of diseases across diverse populations and holds significant potential for future applications across thousands of phenotypes.

## Data availability

Simulation results, EDGE GWAS summary statistics for UK BioBank and meta-analyses are available in supplementary files and tables. The full summary-level association data from the individual population association analyses of MVP data for both phenotypes will be available via the dbGaP study accession phs001672. R codes for data cleaning and plotting and Python codes for simulation construction, α calculation, and α application are available at https://github.com/nicenzhou/EDGE. Research using UK BioBank was conducted under approved project 52374.

## URLs

PLINK v1.9 (https://www.cog-genomics.org/plink/1.9/),

pandas-genomics (BAMS) v0.11.0 (https://github.com/HallLab/pandas-genomics),

CLARITE v2.2.0 (https://clarite-python.readthedocs.io/en/latest/),

PLATO v2.1.0 (https://ritchielab.org/plato),

LDtrait (https://ldlink.nih.gov/?tab=ldtrait),

R statistical software (https://www.R-project.org),

Python Programming Language (https://www.python.org/)

## Acknowledgement

This research is based on data from the Million Veteran Program, Office of Research and Development, Veterans Health Administration, and was supported by Veterans Administration awards I01-01BX003362 (P.S.T.). The content of this manuscript does not represent the views of the Department of Veterans Affairs or the United States Government.

This work is also supported by the USDA National Institute of Food and Agriculture and Hatch Appropriations under Project #PEN04275 and Accession #1018544, startup funds from the College of Agricultural Sciences, Pennsylvania State University (https://agsci.psu.edu/), and the Dr. Frances Keesler Graham Early Career Professorship from the Social Science Research Institute, Pennsylvania State University (https://ssri.psu.edu/) to MAH. The funders had no role in study design, data collection and analysis, decision to publish, or preparation of the manuscript.

We would like to acknowledge Mr. John McGuigan (The Pennsylvania State University) for constructing the pandas-genomics and CLARITE software, Dr. Mudong Zeng (The Pennsylvania State University) and Dr. Zhe Zhang (The University of North Carolina at Chapel Hill) for their advice on statistical model evaluations, Dr. Pik Fang Kho (VA Palo Alto Healthcare System; Stanford University) and Dr. Shoa L. Clarke (VA Palo Alto Healthcare System; Stanford University) for providing the HARE/ancestry and CAD classifications for the MVP cohort, Dr. Tomas Gonzalez Zarzar (Pontificia Universidad Católica de Chile), Dr. Kristin Passero (Virginia Commonwealth University), and Mrs. Nicole Palmiero (University of Pennsylvania) for joining and commenting on EDGE meetings, and Dr. Lin Song (University of Wisconsin-Madison) and Dr. Anye Shi (Cornell University) for their suggestions on manuscript writing and organization.

## Author contributions

Concept and design: J.Z., T.L.A., S.S.V., and M.A.H.

Data acquisition: J.Z., P.S.T., K-M.C., and T.L.A.

Funding Acquisition: P.S.T. and K-M.C.

Data simulation, analysis, and interpretation: J.Z.

Parallel computing for simulation: AL.GR.

Software testing (CLARITE and pandas-genomics): J.Z., AL.GR., and L.G.

Drafting of the manuscript: J.Z.

Revision of the manuscript: All authors

## Supplementary files and tables

Figs. S1 to S15 in File Supplement Figure

Tables. S1 to S10 in Spreadsheet Supplement Table

