## Supplement Figures for "Flexibly encoded genome-wide association study identifies novel nonadditive genetic risk variants for cardiometabolic traits"

**The supplement file includes:**

VA Million Veteran Program: Core Acknowledgements for Publications
Figs. S1 to S15

**VA Million Veteran Program:**
**Core Acknowledgements for Publications**
**May 2024**

### MVP Program Office

- Sumitra Muralidhar, Ph.D., Program Director
  US Department of Veterans Affairs, 810 Vermont Avenue NW, Washington, DC 20420
- Jennifer Moser, Ph.D., Associate Director, Scientific Programs
  US Department of Veterans Affairs, 810 Vermont Avenue NW, Washington, DC 20420
- Jennifer E. Deen, B.S., Associate Director, Cohort & Public Relations
  US Department of Veterans Affairs, 810 Vermont Avenue NW, Washington, DC 20420

### MVP Executive Committee

- Co-Chair: Philip S. Tsao, Ph.D.
  VA Palo Alto Health Care System, 3801 Miranda Avenue, Palo Alto, CA 94304
- Co-Chair: Sumitra Muralidhar, Ph.D.
  US Department of Veterans Affairs, 810 Vermont Avenue NW, Washington, DC 20420
- J. Michael Gaziano, M.D., M.P.H.
  VA Boston Healthcare System, 150 S. Huntington Avenue, Boston, MA 02130
- Elizabeth Hauser, Ph.D.
  Durham VA Medical Center, 508 Fulton Street, Durham, NC 27705
- Amy Kilbourne, Ph.D., M.P.H.
  VA HSR&D, 2215 Fuller Road, Ann Arbor, MI 48105
- Michael Matheny, M.D., M.S., M.P.H.
  VA Tennessee Valley Healthcare System, 1310 24th Ave. South, Nashville, TN 37212
- Dave Oslin, M.D.
  Philadelphia VA Medical Center, 3900 Woodland Avenue, Philadelphia, PA 19104
- Deepak Voora, MD
  Durham VA Medical Center, 508 Fulton Street, Durham, NC 27705

### MVP Co-Principal Investigators

- J. Michael Gaziano, M.D., M.P.H.
  VA Boston Healthcare System, 150 S. Huntington Avenue, Boston, MA 02130
- Philip S. Tsao, Ph.D.
  VA Palo Alto Health Care System, 3801 Miranda Avenue, Palo Alto, CA 94304

### MVP Core Operations

- Jessica V. Brewer, M.P.H., Director, MVP Cohort Operations
  VA Boston Healthcare System, 150 S. Huntington Avenue, Boston, MA 02130
- Mary T. Brophy M.D., M.P.H., Director, VA Central Biorepository
  VA Boston Healthcare System, 150 S. Huntington Avenue, Boston, MA 02130
- Kelly Cho, M.P.H, Ph.D., Director, MVP Phenomics

VA Boston Healthcare System, 150 S. Huntington Avenue, Boston, MA 02130

- Lori Churby, B.S., Director, MVP Regulatory Affairs
  VA Palo Alto Health Care System, 3801 Miranda Avenue, Palo Alto, CA 94304
- Scott L. DuVall, Ph.D., Director, VA Informatics and Computing Infrastructure (VINCI)
  VA Salt Lake City Health Care System, 500 Foothill Drive, Salt Lake City, UT 84148
- Saiju Pyarajan Ph.D., Director, Data and Computational Sciences
  VA Boston Healthcare System, 150 S. Huntington Avenue, Boston, MA 02130
- Robert Ringer, Pharm.D., Director, VA Albuquerque Central Biorepository
  New Mexico VA Health Care System, 1501 San Pedro Drive SE, Albuquerque, NM 87108
- Luis E. Selva, Ph.D., Director, MVP Biorepository Coordination
  VA Boston Healthcare System, 150 S. Huntington Avenue, Boston, MA 02130
- Shahpoor (Alex) Shayan, M.S., Director, MVP PRE Informatics
  VA Boston Healthcare System, 150 S. Huntington Avenue, Boston, MA 02130
- Brady Stephens, M.S., Principal Investigator, MVP Information Center
  Canandaigua VA Medical Center, 400 Fort Hill Avenue, Canandaigua, NY 14424
- Stacey B. Whitbourne, Ph.D., Director, MVP Cohort Development and Management
  VA Boston Healthcare System, 150 S. Huntington Avenue, Boston, MA 02130

**MVP Publications and Presentations Committee**

- Co-Chair: Themistocles L. Assimes, M.D., Ph. D
  VA Palo Alto Health Care System, 3801 Miranda Avenue, Palo Alto, CA 94304
- Co-Chair: Adriana Hung, M.D.; M.P.H
  VA Tennessee Valley Healthcare System, 1310 24$^{th}$ Ave. South, Nashville, TN 37212
- Co-Chair: Henry Kranzler, M.D.
  Philadelphia VA Medical Center, 3900 Woodland Avenue, Philadelphia, PA 19104

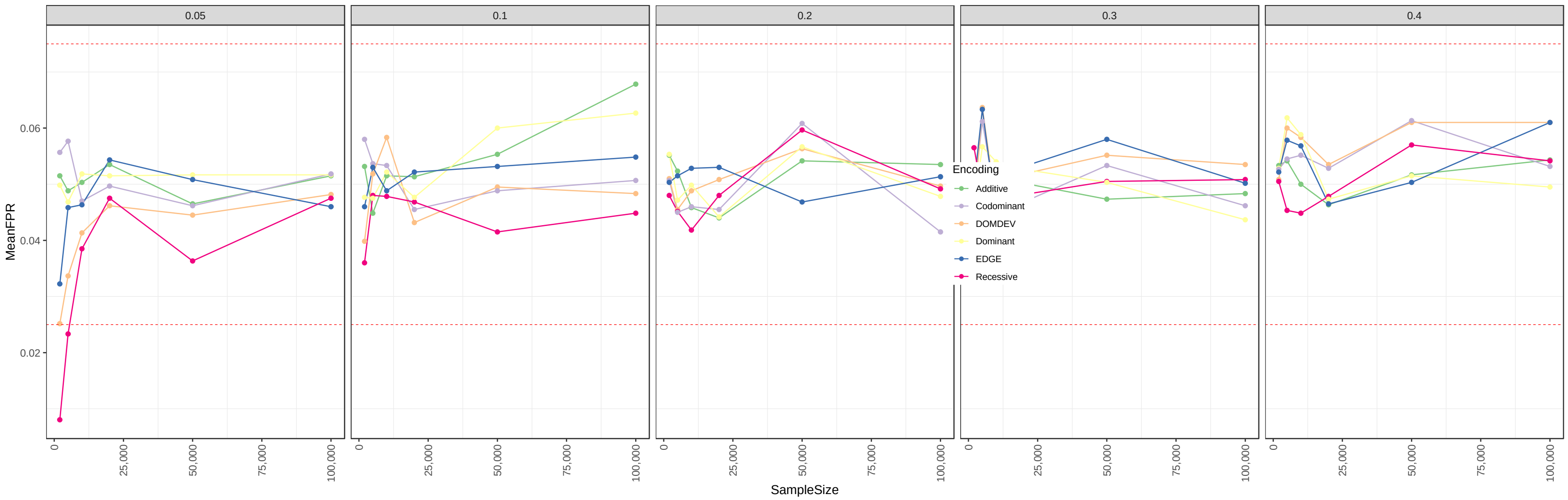


**Figure S1.** Inflation of using EDGE and other five encodings. The dashed line represents Bradley's liberal criteria of 2.5%-7.5%

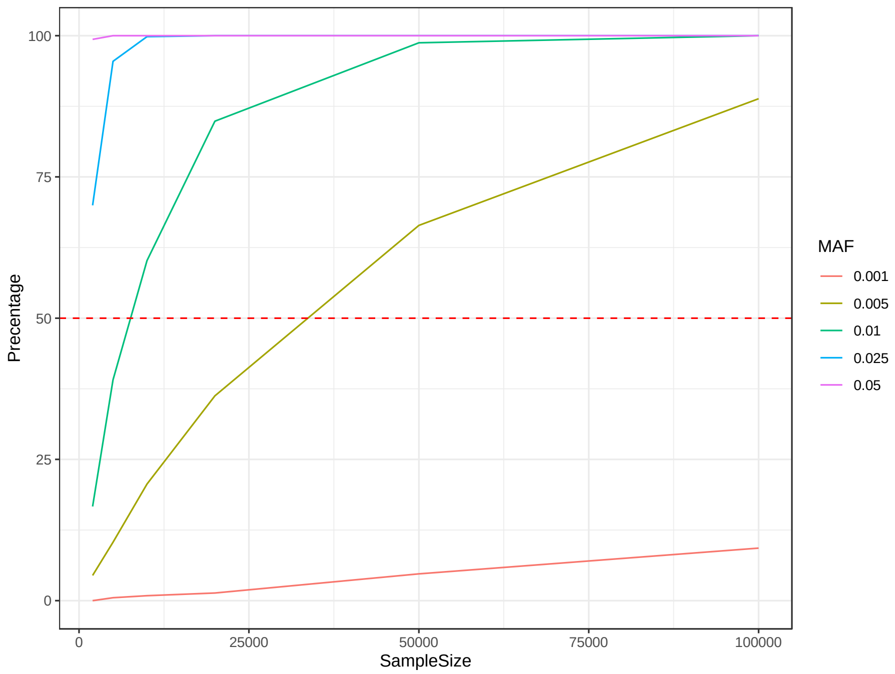


**Figure S2.** Convergence of using EDGE towards to the rare variants.

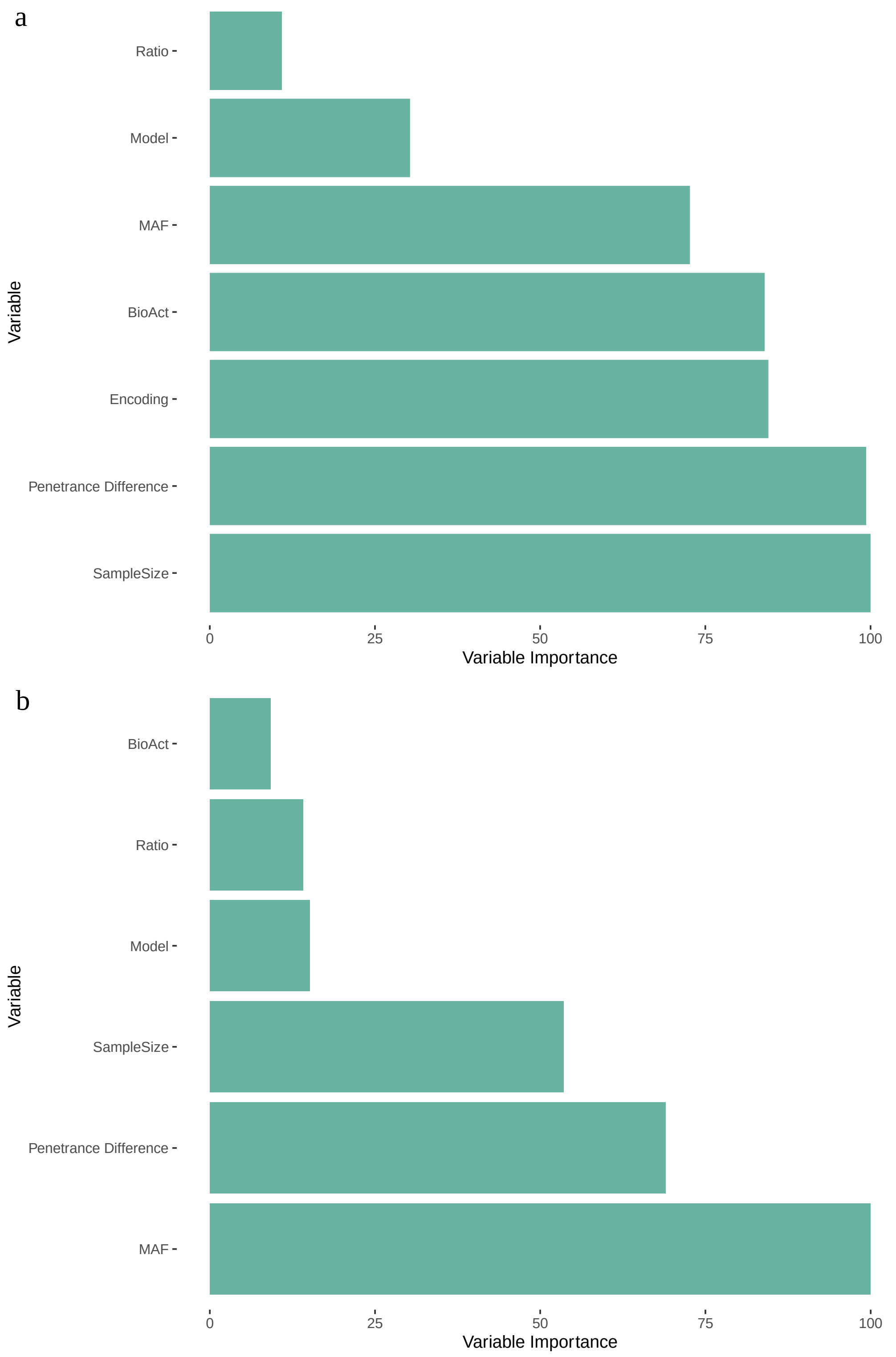


**Figure S3.** Ranked variable importance for a) power and b) alpha calculations through an overfitted random forest analyses. All the variable importance is scaled by considering the largest one as the 100%. MAF: minor allele frequency; BioAct: Inheritance models.

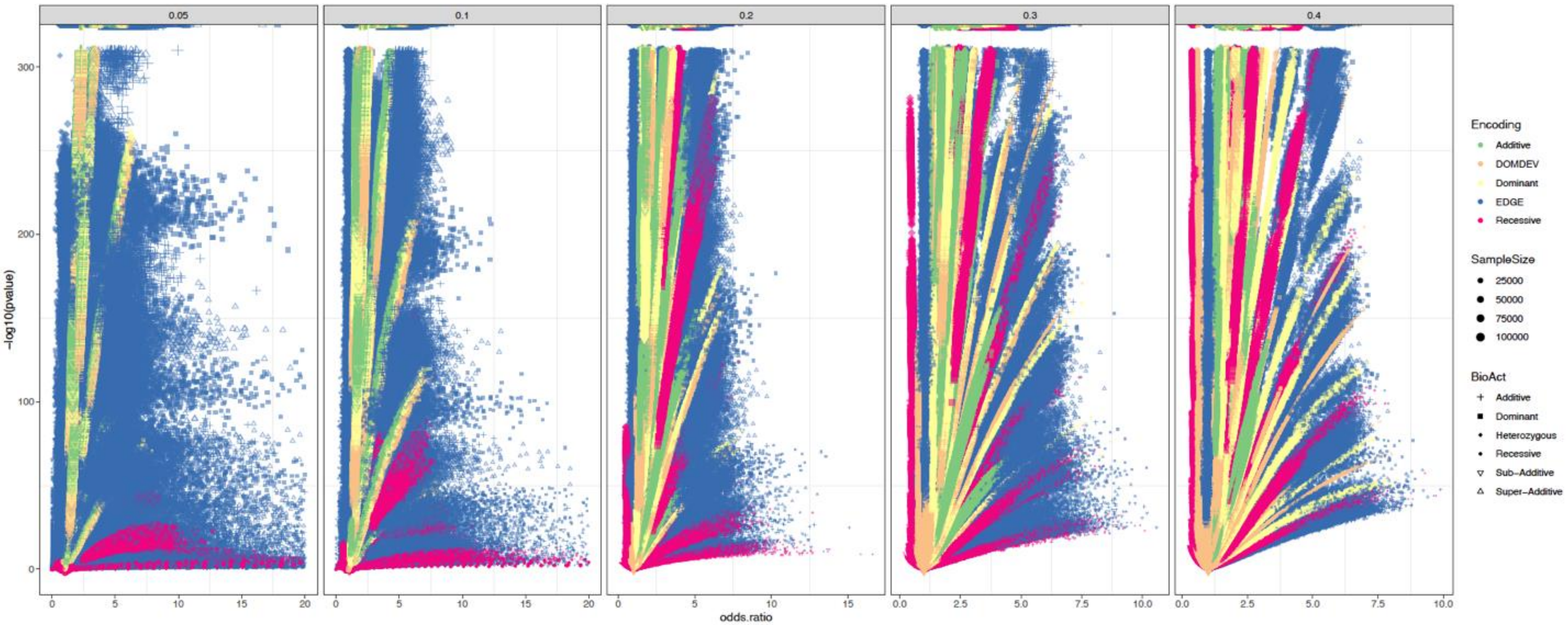


**Figure S4.** Volcano plots for showing the distribution of odds ratio and p-values.

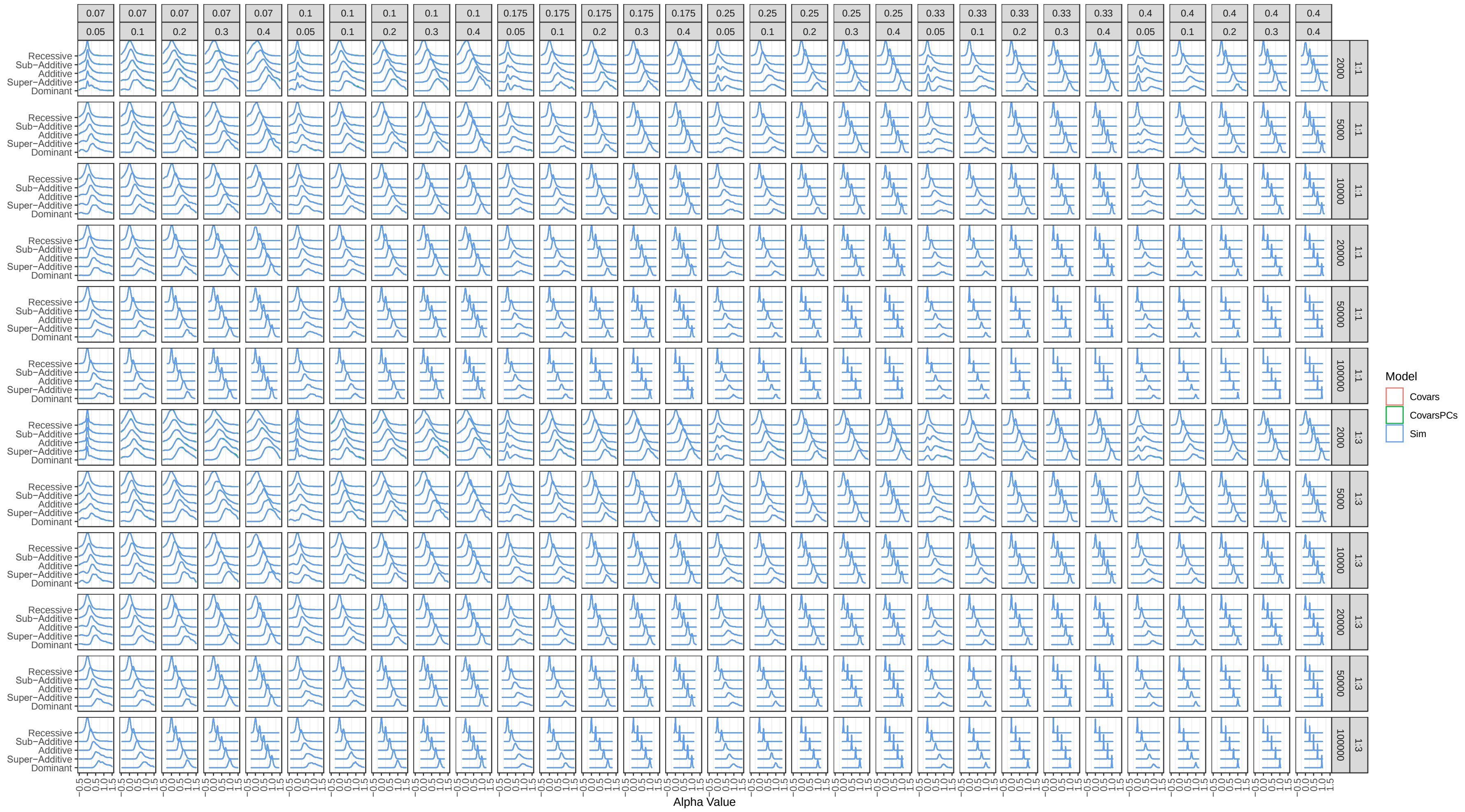


**Figure S5.** Distribution of the alpha values under different settings of simulations.

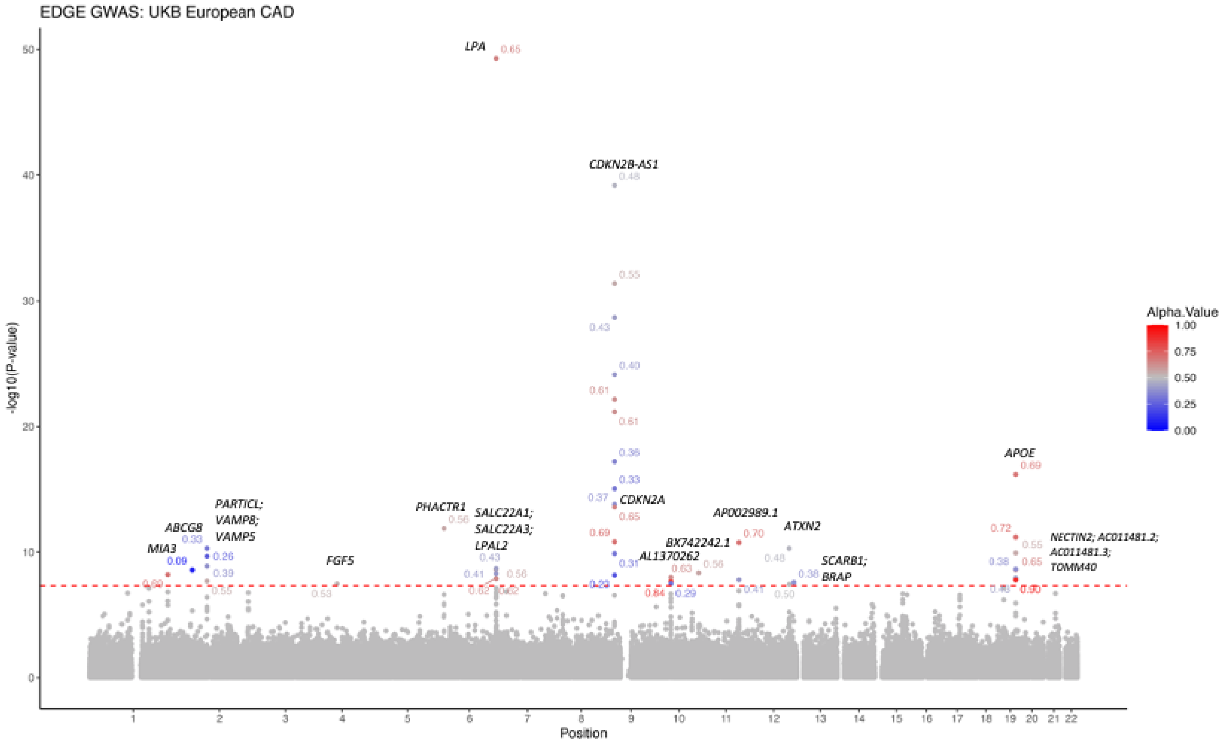


**Figure S6.** GWAS for CAD in UK Biobank European ancestry group of using EDGE encoding. The red dashed lines represent the genome-wide significance at $5\times10^{-8}$.

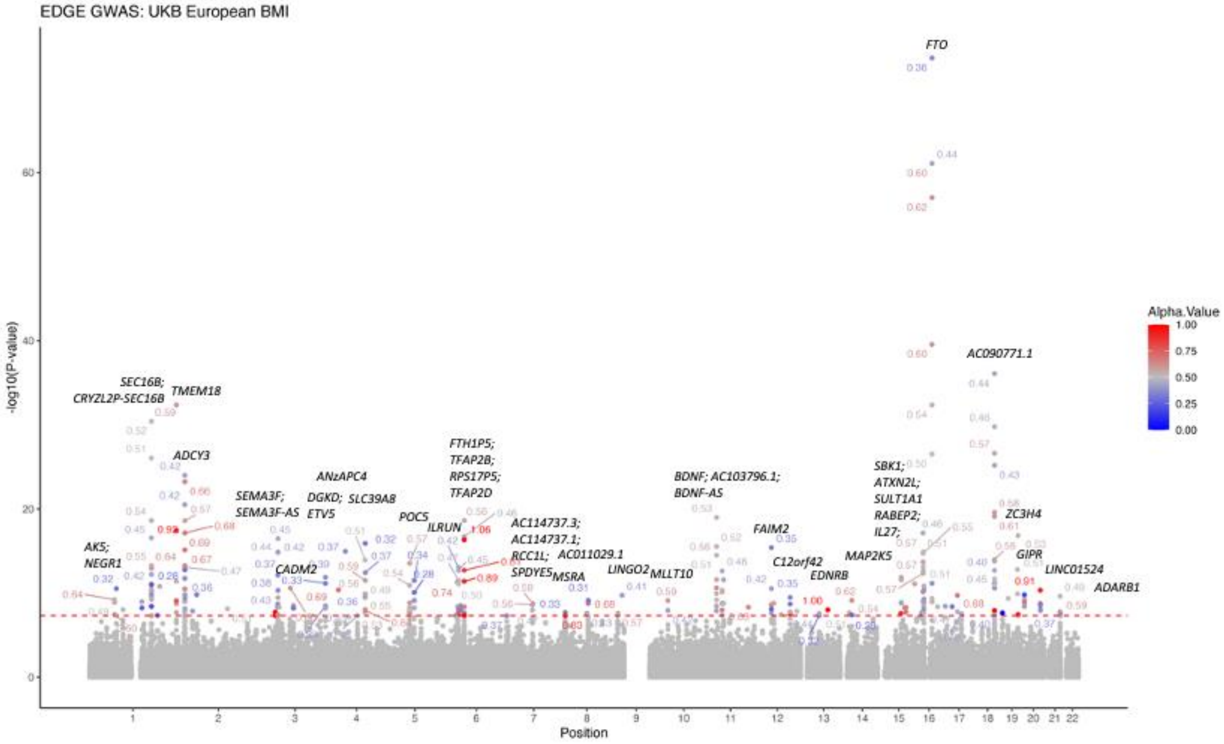


**Figure S7.** GWAS for BMI in UK Biobank European ancestry group of using EDGE encoding. The red dashed lines represent the genome-wide significance at $5\times10^{-8}$.

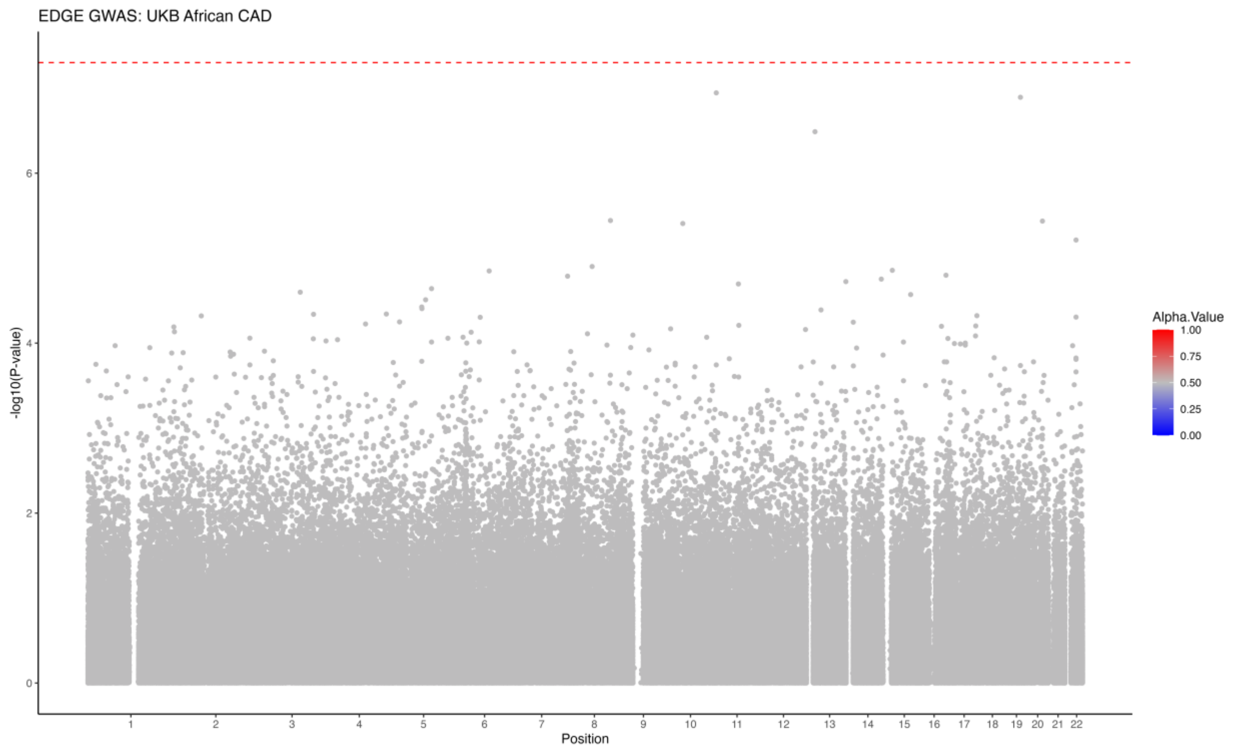


**Figure S8.** GWAS for CAD in UK Biobank African ancestry group of using EDGE encoding. The red dashed lines represent the genome-wide significance at $5\times10^{-8}$.

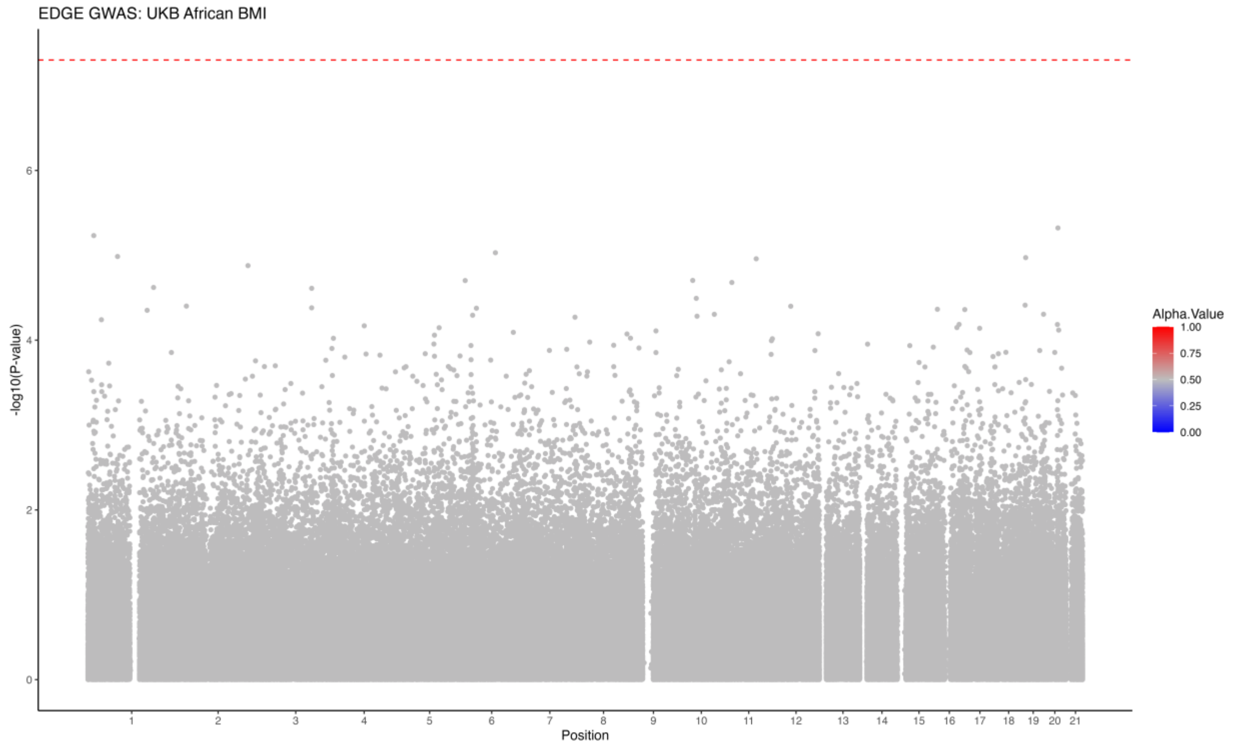


**Figure S9.** GWAS for BMI in UK Biobank African ancestry group of using EDGE encoding. The red dashed lines represent the genome-wide significance at $5\times10^{-8}$.

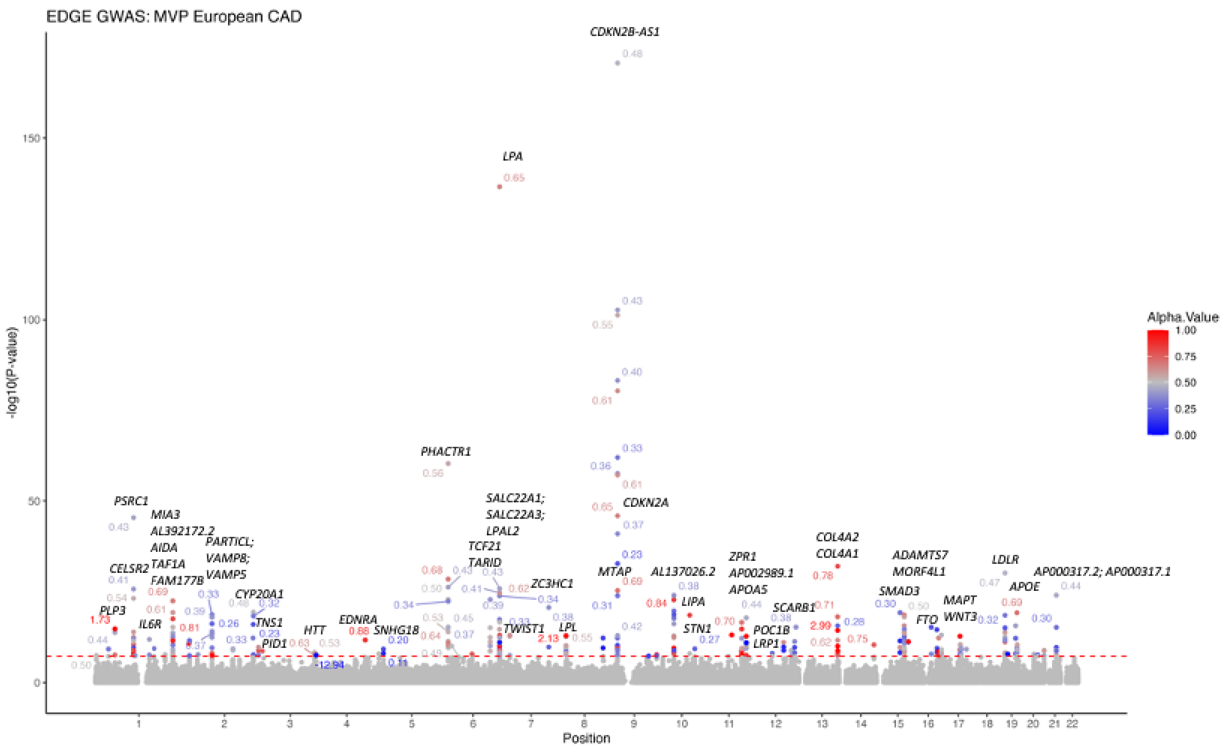


**Figure S10.** GWAS for CAD in MVP European ancestry group of using EDGE encoding. The red dashed lines represent the genome-wide significance at $5{\times}10^{-8}$.

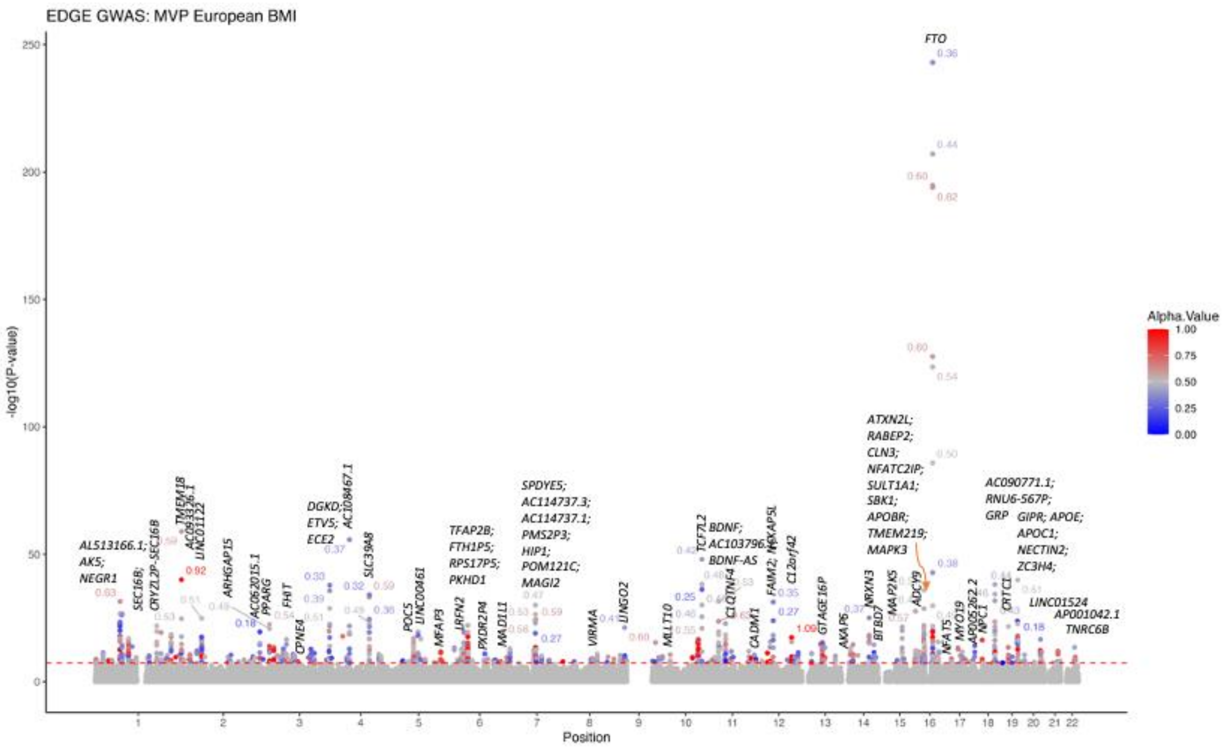


**Figure S11.** GWAS for BMI in MVP European ancestry group of using EDGE encoding. The red dashed lines represent the genome-wide significance at $5{\times}10^{-8}$.

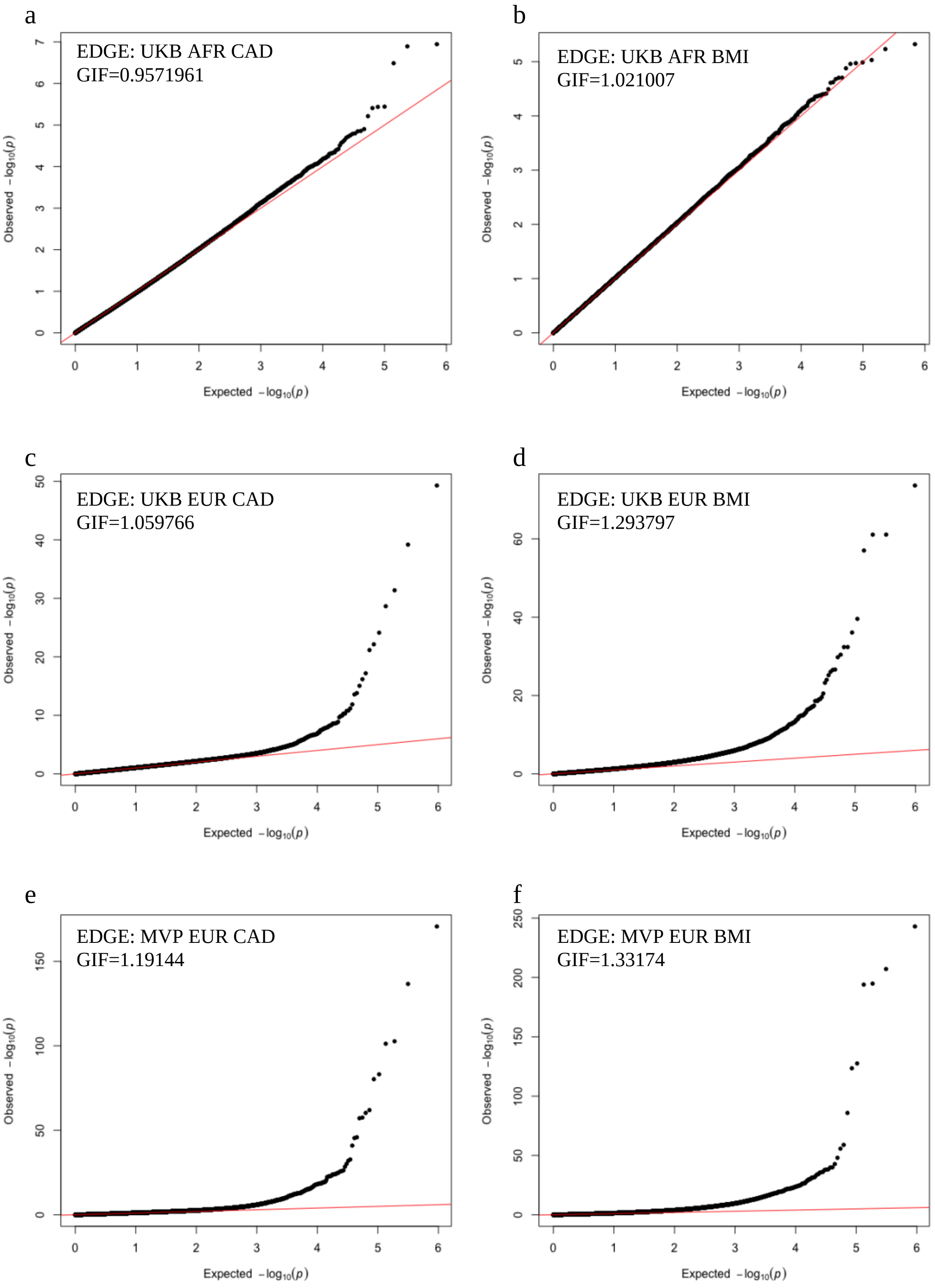


**Figure S12.** Q-Q plot for EDGE GWAS with a) UK Biobank AFR CAD, b) UK Biobank AFR BMI, c) UK Biobank EUR CAD, d) UK Biobank EUR BMI, e) MVP EUR CAD, and f) MVP EUR BMI.

a

b

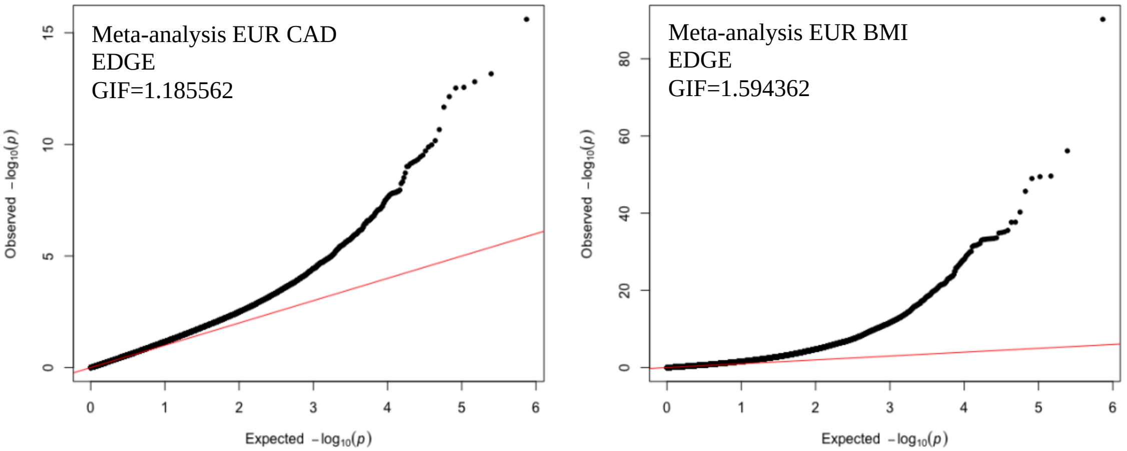


**Figure S13.** Q-Q plot for meta-analysis among SNPs with $I^2 \leq 40$ for EUR from UK Biobank and MVP using a) EDGE for CAD, and b) EDGE for BMI.

a

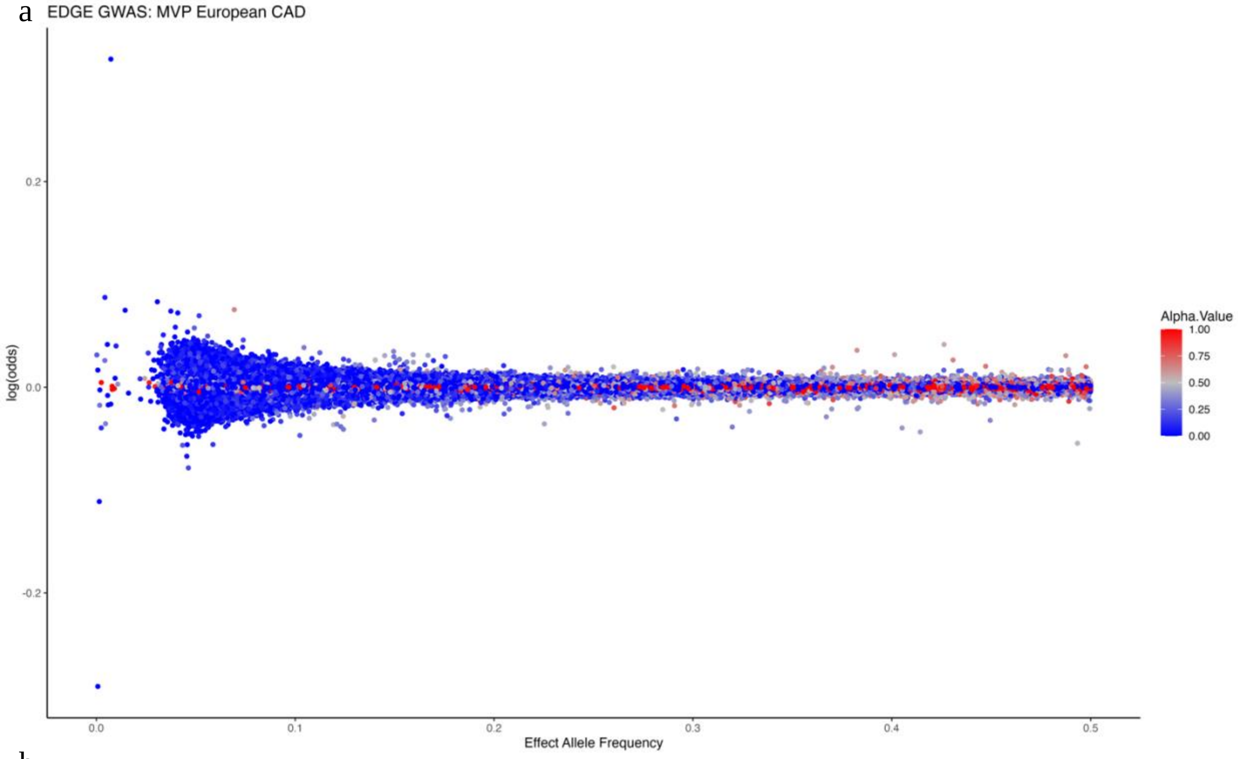

EDGE GWAS: MVP European CAD
log(odds)
Effect Allele Frequency
Alpha.Value
1.00
0.75
0.50
0.25
0.00


b

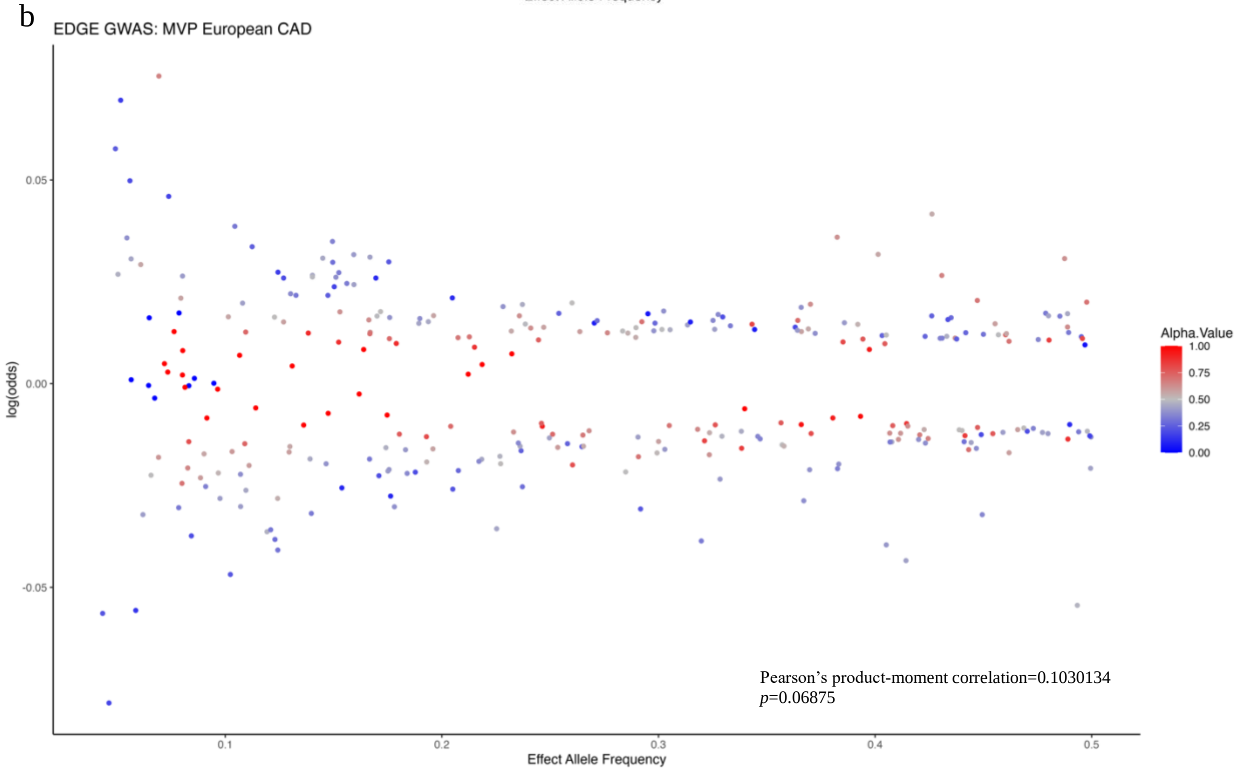

EDGE GWAS: MVP European CAD
log(odds)
Effect Allele Frequency
Alpha.Value
1.00
0.75
0.50
0.25
0.00
Pearson's product-moment correlation=0.1030134
p=0.06875

c

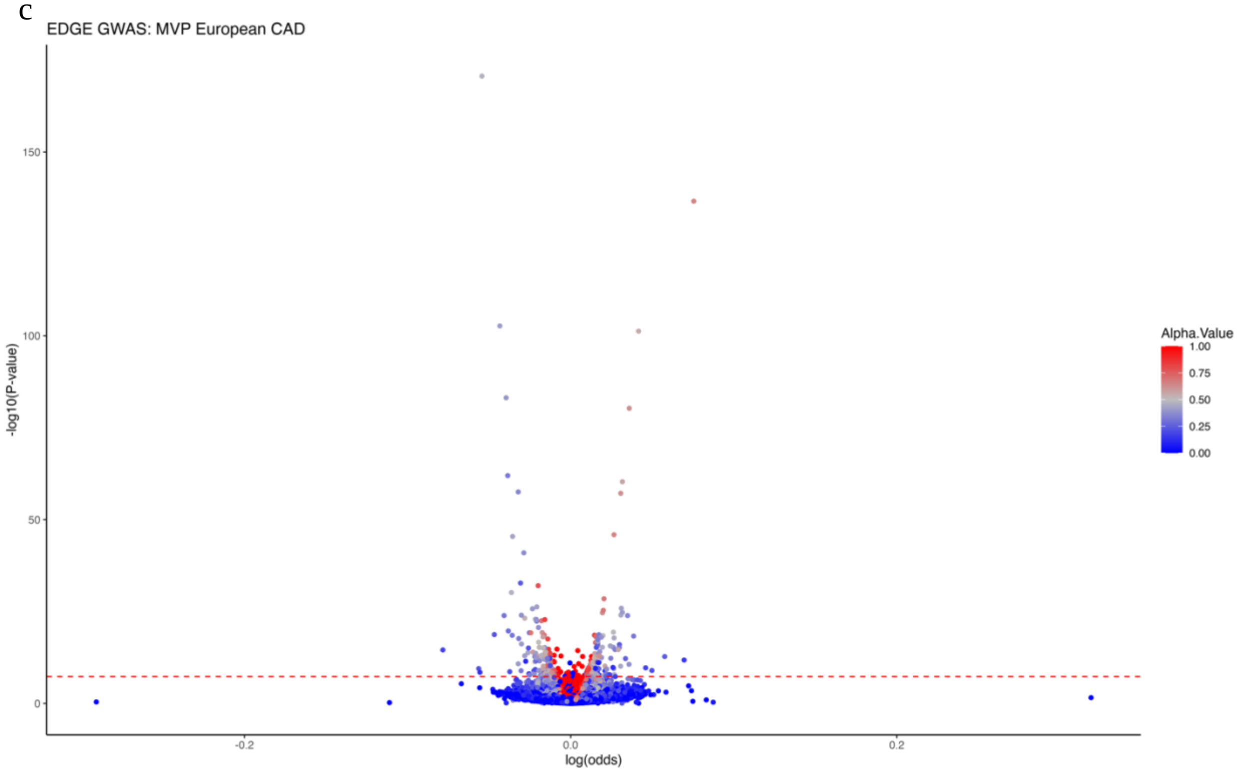


**Figure S14.** The distribution of the a) log(odds) and EAF, b) log(odds) and EAF for significant results only, and c) -log10(p-value) and log(odds) for MVP EUR CAD. The red dashed lines represent the genome-wide significance (GWS) at $5\times10^{-8}$. The Pearson's product-moment correlation was calculated between the absolute log(odds) and alpha values for variants with GWS based on MVP results.

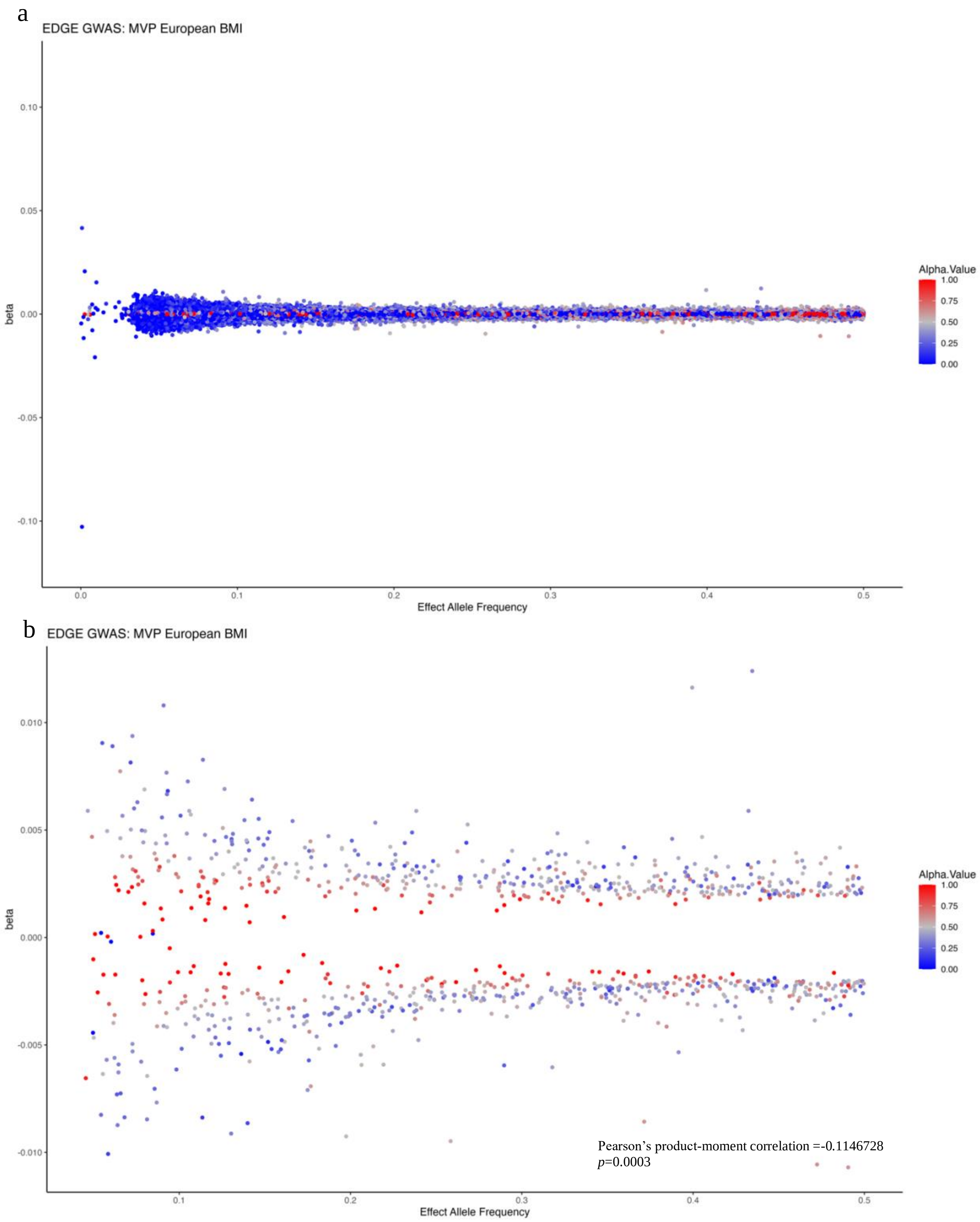
a
EDGE GWAS: MVP European BMI
beta
Effect Allele Frequency
Alpha.Value
b
EDGE GWAS: MVP European BMI
beta
Effect Allele Frequency
Alpha.Value
Pearson's product-moment correlation =-0.1146728
p=0.0003

c

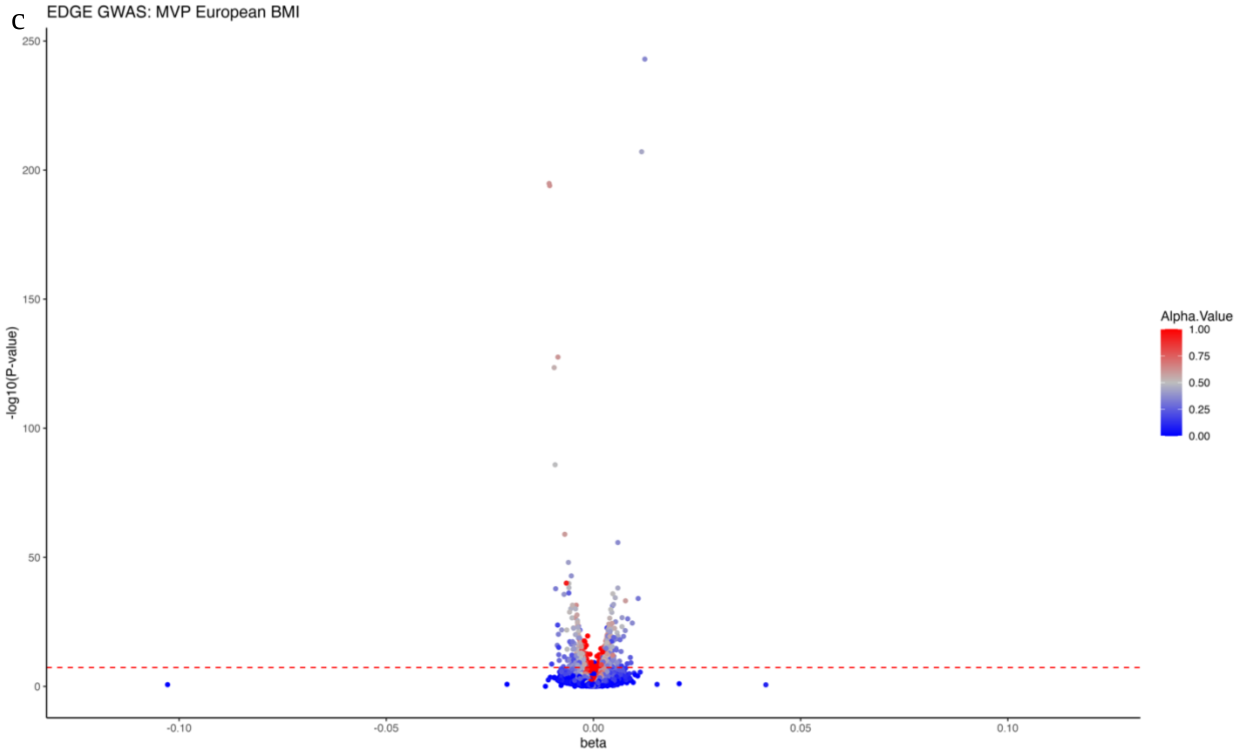


**Figure S15.** The distribution of the a) beta and EAF, b) beta and EAF for significant results only, and c) -log10(p-value) and beta for MVP EUR BMI. The red dashed lines represent the genome-wide significance (GWS) at $5\times10^{-8}$. The Pearson's product-moment correlation was calculated between the absolute beta coefficient and alpha values for variants with GWS based on MVP results.
